# Does LLM Assistance Improve Healthcare Delivery? An Evaluation Using On-site Physicians and Laboratory Tests∗

**DOI:** 10.1101/2025.10.31.25339278

**Authors:** Jason Abaluck, Robert Pless, Nirmal Ravi, Anja Sautmann, Aaron Schwartz

## Abstract

We deployed large language model (LLM) decision support for health workers at two outpatient clinics in Nigeria. For each patient, health workers drafted care plans that were optionally revised after LLM feedback. We compared unassisted and assisted plans using blinded randomized assessments by on-site physicians who evaluated and treated the same patients and using results from laboratory tests for common conditions. Academic physicians performed blinded retrospective reviews of a subset of notes. In response to LLM feedback, health workers changed their prescribing for more than half of patients. Health workers reported high satisfaction with LLM feedback and retrospective academic reviewers rated LLM-assisted plans more favorably. However, on-site physicians observed little to no improvement in diagnostic alignment or treatment decisions. Laboratory testing showed mixed effects of LLM-assistance, which removed negative tests for malaria but added them for urinary tract infection and anemia, with no significant increase in the detection rates for the tested conditions. This highlights a gap between chart-based reviews and real-world clinical relevance that may be especially important in evaluating the effectiveness of LLM-based interventions.

## 1 Introduction

Large language models (LLMs) have the potential to improve health provider decision-making without requiring substantial infrastructure to deploy (Pressman et al., 2024). LLMs have shown diagnostic performance comparable to general physicians on written tests and in simulated patient encounters such as vignettes or model patients (Takita et al., 2025; Huang et al., 2024; Tu et al., 2025). A smaller number of studies have tested LLMs with real patient information, or LLM decision support in realistic clinical environment (Goh et al., 2024; Cadamuro et al., 2023; Gaber et al., 2025; Hager et al., 2024). Evaluation of LLM performance is typically retrospective, comparing the LLM’s conclusions with the decisions of the treating physician (Shmilovitch et al., 2025; Yıldırım et al., 2025). We are only aware of one peer-reviewed and one unpublished study that implement forms of LLM support in a live clinic setting (Korom et al., 2025; Wan et al., 2024). In both, the evaluation of quality of care with and without LLM is done via retrospective review, which by its nature, focuses on the internal consistency of the patient record and cannot account for information that the treating provider may have missed or decided not to record.

We build on the existing literature by evaluating LLM-assisted care using blinded review by physicians who independently assessed the same patient, as well as objective medical test results. We use these approaches and others to evaluate a prototype intervention in two outpatient clinics in Kano, Nigeria, in which an LLM gives providers an instant “second opinion” on their care plans. This intervention models lightweight approaches that could be broadly deployed, and mirrors the informal ways providers are already consulting LLMs (Blease et al., 2024). Health workers (here Community Health Extension Workers, or CHEWs) drafted initial, unassisted care plans and then revised them after receiving LLM feedback, producing paired unassisted and assisted notes. All patients were subsequently examined by on-site physicians (Medical Officers, or MOs), who developed their own care plans, treated and discharged the patients (to avoid any impacts of LLM use on care received), and then evaluated both health worker plans in a blinded manner, using their direct, personal knowledge of the patient.

Our design allows us to compare unassisted and LLM-assisted care plans using four complementary lenses: (1) subjective ratings from on-site physicians who saw the same patients; (2) concordance between the health worker and on-site physician care plans; (3) agreement of testing and treatment decisions with laboratory test results for the three most commonly tested conditions (malaria, urinary tract infection, and anemia); and (4) blinded retrospective assessment of a subsample of patients by third-party academic physicians (MDs) who reviewed the written notes produced by the health workers and the physician who saw the patient.

In all four domains, health workers’ care plans *prior* to LLM assistance show substantial deficits. We find that LLM assistance led health workers to meaningfully revise diagnoses (41% of notes), test ordering (33% of notes), and prescribing decisions (54% of notes). The health workers themselves overwhelmingly reported that they found the LLM feedback helpful. Academic physicians also rated LLM-assisted notes more favorably in retrospective review. However, on-site physicians who had examined the same patients observed little or no improvement in diagnostic alignment or treatment decisions, and assisted plans did not resemble physician plans more closely. Comparison with laboratory tests revealed mixed effects: the LLM led the health workers to remove negative tests for malaria, but added them for urinary tract infection and anemia, and did not encourage testing for cases later found to be positive for the condition. There was subsequently no improvement in the appropriateness of treatment given conditions detected by testing.

When analyzing the LLM feedback itself, we find that the LLM makes 3.75 recommendations per patient; these recommendations were about equally likely to increase or decrease alignment with physician testing decisions and prescriptions, although they better aligned with physician advice. Health workers followed about one third of recommendations, but were only slightly more likely than chance to follow recommendations that improved physician concordance; in other words, the “human in the loop” does not serve to filter out the less helpful LLM advice. When presented with the symptoms noted by the on-site physician, LLM feedback improves, but only enough to close a small fraction of the gap between health worker and physician plans.

These findings reveal a notable divergence: LLM feedback improves plan documentation according to retrospective academic reviewers, yet fails to produce consistent improvements according to physicians with firsthand knowledge of the patient, or according to laboratory results. These findings suggest a potential disconnect between retrospective evaluation and locally observed clinical performance.

## 2 Study Design

### 2.1 Experimental Design

#### Setting

This study was conducted in two private clinics in the city of Kano in Kano State, operated by EHA Clinics Nigeria. One is a community clinic staffed primarily by Community Health Extension Workers (CHEWs), where patients are typically seen by CHEWs. The other is a full-service clinic where patients are typically seen by Medical Officers. The full-service clinic has a pharmacy and an in-house laboratory for routine laboratory tests, which is also used by the community clinic. For this study, all enrolled patients at both clinics were seen first by a CHEW and then by an on-site MO, regardless of the clinic’s standard staffing model.

Community Health Extension Workers in Nigeria undergo a three-year licensed diploma program. Their training focuses on community outreach, basic preventive and curative care, maternal health, and referrals to higher-level providers (Ajisegiri et al., 2023; Dotimi et al., 2024). While they generally work under the supervision of a nurse or MO, CHEWs may be *de facto* the primary provider in a public clinic or health outpost in rural or underserved areas (Okoroafor et al., 2022). In contrast, Medical Officers have completed training that includes at minimum a four-year medical bachelor degree and internship; their scope of practice is that of a full physician. In a large randomized trial, Okeke (2023) showed that simply deploying one additional MO to a public clinic in Nigeria significantly improved quality of care and patient outcomes.

EHA Clinics provides training for CHEWs and MOs in their clinical protocols, which include rules to prevent the overuse of antimalarials and antibiotics. EHA Clinics also uses a custom electronic medical record (EMR) system to record patient information and track medical test orders and prescriptions. For this study, the information recorded in the EMR was used in the background to collate patient information collected by the provider into a SOAP note ((S)ubjective-(O)bjective-(A)ssessment-(P)lan).

#### Intervention and patient flow

Patients were enrolled and informed consent was obtained before they were accompanied through the steps of the study by dedicated research staff. A patient’s first clinician encounter was with a CHEW, who conducted the patient consultation and prepared a “conditional” SOAP note that specified the provisional diagnosis and prescriptions conditional on laboratory test results.^1^

The health worker then submitted the “unassisted” SOAP note to the LLM indirectly through the EMR for feedback. The LLM prompt and sample LLM feedback are shown in Appendix A. The LLM prompt was developed in extensive piloting and review of simulated feedback and was in particular aimed at producing concise responses that did not rely on excessive laboratory testing (McPeak et al., 2024). Upon receiving the feedback from the LLM in the EMR, the health worker prepared an updated “assisted” (conditional) SOAP note.

It is important to note that health workers did not actually order any medical tests or diagnose and treat patients in this study for ethical reasons. They were trained to conduct a patient consultation that is as close to their daily practice as possible, but to refrain from any diagnosis or remarks on the patient’s health to the patient. They were also trained to view the LLM feedback as potentially flawed and to only adopt advice that they agreed with. It is also important to note that the feedback from the LLM was never used for any treatment or diagnostic purposes.

Patients next saw the physician (MO) on site who conducted an independent evaluation. Patients were advised upon enrollment to treat the two consultations as independent and repeat information to the physician even if they had shared it with the health worker previously. The physician prepared their own conditional SOAP note and ordered any laboratory testing needed to diagnose the patient. Test results that were available immediately were incorporated into the physician’s patient record and included in the discharge SOAP note. We interpret this discharge note as illustrating a typical care plan that would have been recommended, had the patient visited a physician at these clinics outside of the study.

At this stage of the study, patients were re-approached by research staff who offered additional point-of-care laboratory testing for three common conditions – malaria, anemia, and urinary tract infection – based on a set of demographic criteria and symptom checks (see below and Appendix C.2 for details). The purpose of this testing step was to collect additional objective data beyond the results of physician-ordered tests so that the study could better evaluate the quality of health worker care plans. Patients were unaware up to this point that they could be offered these three tests, and the test results were not made available at the time of the clinic visit. Rather, blood and urine samples were collected and patients were discharged, with any supplemental testing conducted in the course of the next 1-2 days. This design was chosen to avoid contamination of the physician’s testing decisions. As the final step, typically in the evening or on the next day, the on-site physician reviewed the full patient record including any additional test results from the research staff, updated their own SOAP note (and flagged patients with actionable findings for follow-up), and then carried out a blinded evaluation of the unassisted and assisted health worker notes, labeled randomly SOAP A or SOAP B. By design, some “missing” or “incorrect” elements in the MO evaluation may reflect information gaps rather than clinical mis-reasoning. This likely inflates baseline error rates compared to evaluations where both providers have equal information. However, because both unassisted and assisted health worker notes were drafted under identical information constraints, the estimated LLM effect remains unbiased. The physician evaluation of the notes measures whether LLM feedback helps health workers approximate the decisions of a fully informed physician based on clinical presentation alone, which reflects a realistic and clinically relevant scenario for decision support tools.

#### Sample

Study funding was projected to cover approximately 500 patients in the main sample. After extensive piloting, the LLM-assistance intervention was initially rolled out on January 30, 2025, and study patient encounters were conducted through May 26. Due to an error in the EMR system, randomization of care plan labels was initially not implemented correctly, and physicians always reviewed the LLM-unassisted note first (labeled SOAP A). This error was corrected by February 25. Thus, while we include the full sample of 660 patient visits in analyses of objective outcome measures (i.e. direct comparisons of care plans, or comparison of care plans to medical test results, etc..), we use only the subsample of 491 patient encounters after February 25 in any analyses of the physicians’ evaluation of health worker notes (this was preregistered at the time when the error was discovered).

Table 1 gives an overview of the two samples. This population extends beyond the intended scope of a health worker’s practice, but reflects patients these health workers may treat in understaffed public clinics.

**Table 1:** Study Samples: All Patients and Fully Randomized Sample.

|  | Full sample<br>Mean | Blinded sample<br>Mean |
| --- | --- | --- |
| Share Female | 0.64 | 0.66 |
| Share Age 0-4 | 0.06 | 0.07 |
| Share Age 5-14 | 0.11 | 0.13 |
| Share Age 15-45 | 0.72 | 0.71 |
| Share community clinic | 0.49 | 0.48 |
| Share health worker would refer to physician | 0.77 | 0.77 |
| Share physician rated seriously ill | 0.33 | 0.24 |
| Share unassisted note is note A | 0.62 | 0.49 |
| N (Patients) | 660 | 491 |
| N (Health workers) | 21 | 21 |
| N (Physicians) | 8 | 6 |

This table reports demographic and clinical characteristics for two samples. The full sample includes all patient visits from January 30 to May 26, 2025, representing the complete study period. The fully randomized sample restricts to visits from February 25 to May 26, 2025, when the order of health worker notes presented to physicians was correctly randomized. All statistics report means with standard deviations in parentheses. “Share unassisted note is note A” indicates the proportion of cases where the unassisted health worker note was randomly assigned to be presented first to the physician.

The demographic profile is broad, with a significant share of adolescents and adult men who are not a focus of community health interventions; 24% of patients (33% in the larger sample) are judged by the physician to be seriously ill; and in 77% of the cases the health worker states that outside of the study they would refer this patient to a physician.^2^ It is worth noting that EHA Clinics have higher staffing levels than typical public clinics in Nigeria. For example, a physician is available through telemedicine at all times in the community clinic. Physicians are also the main providers at the full-service clinic. These factors likely influence patient composition.

The second-to-last panel verifies the balance in randomized assignment in the sample after February 25. The intervention was carried out by 21 health workers who rotated through the study and each saw between 25 and 40 patients; and there were a total of 8 reviewing physicians.

### 2.2 Data Sources

We use two main sources of data; patient EMR data and survey data from health workers (CHEWs), physicians (MOs), and third-party academic physicians (MDs).

#### 2.2.1 Patient encounter data in the EMR

All patient data entered by the intake nurse, the health worker and physician, and the lab technician are saved in the EMR. As described above, a total of five SOAP notes were created for each patient, the LLM-unassisted and LLM-assisted notes by the health worker, the physician’s conditional note (prior to obtaining any lab test results), the discharge note (after seeing the results of lab tests ordered by the physician), and the final physician note which includes any additional test results for malaria, UTI and anemia collected by the study staff. In addition to the text files containing the structured SOAP notes, we collected the values of the individual EMR entry fields within the SOAP notes, such as test results, diagnoses, prescriptions, vital signs, and basic demographic information.

The subjective information in the SOAP notes includes the self-reported chief concern, history of presenting illness, any past medical history, current medications, allergies, and relevant family or social history. Objective data include vital signs, physical examination findings, and lab test results. The assessment and plan contain the diagnosis and differential diagnosis, testing or treatment the clinician is pursuing, including non-prescription advice (e.g., “patient counseled to drink plenty of fluids”), referrals to other clinicians or sites, or warning signs that would warrant another clinic visit.

For each recommended medication order entered into the EMR, the provider must supply detailed dosing information along with a clinical indication, i.e., the related diagnosis. For this study, clinicians could also optionally choose to attach a condition to the prescription based on medical tests ordered. For example, the health worker might use the lab testing section of the EMR to request a malaria test, and then in the prescription section choose a malaria drug that is only given on condition of a positive test result. In standard practice, this conditioning would not be explicitly recorded, but rather a prescription would be entered only if the malaria diagnosis was confirmed by a positive test.

##### Medical test results

For all medical tests conducted, we summarized the physician’s interpretation of the test as “normal” or “abnormal”. As described earlier, we conducted broad testing among our patient sample for three common conditions – malaria, urinary tract infection (UTI), and anemia. Research staff checked a broad list of symptoms intended to identify anyone who should plausibly be tested for one of these three conditions. Patients were then consented for any tests that were not already ordered by the physician. We restricted testing to the most relevant demographic groups-all symptomatic patients for malaria, symptomatic female patients age 7 or older for UTI, and symptomatic patients over 18 for anemia. These test results provide “ground truth” data on the patient’s true disease status. Appendix C.2 provides further detail.

#### 2.2.2 Physician and MD Evaluation of Care Plans

Key measures of the quality of the health worker’s care plans are blinded evaluations by 1) the physician who treated the same patient and 2) a panel of three Medical Doctors (MDs). These MDs work at Bayero University and Aminu Kano Teaching Hospital in Kano and have completed four years of post-graduate training in family and community medicine. They were provided retrospective access to both health worker notes (blinded to LLM-assistance status) and the physician note.

The evaluation was done via a detailed survey in conjunction with (for the physicians) a custom review mask implemented directly in the EMR. The physicians first reviewed any lab test results ordered by the research staff (malaria, UTI, anemia), and updated their own SOAP note to a final SOAP note, and then began review with SOAP note A (randomly selected to be either the unassisted or assisted health worker note). At this stage, they were unable to see SOAP note B.

The physician evaluation focuses on treatment errors in the SOAP note that could cause some form of (medical) harm to the patient. For each SOAP note, the physician first takes stock of qualitative differences between the health worker’s plan and their own plan, then assesses the type of harms that would arise for the patient from any errors identified, and finally rates the SOAP note on a scale for “healthy time lost”.

##### Measuring Healthy Time Lost in DALY

We constructed an anchored scale to measure how much healthy time would have been lost if a health worker plan had been implemented instead of the physician’s plan. The scale translates specific types of clinical error into units of disability-adjusted life year (DALYs), which reflect both length and quality of life. Losing one DALY is equivalent to losing a year of life in perfect health, or two years of life experienced in an unhealthy state with a utility weight of 0.5, etc.

The anchored scale consists of a set of cut-off points that were calibrated using benchmarks of expected DALY loss for clinical mistakes relevant to local medical practice. DALY benchmarks were derived from previously published DALY or QALY estimates from the medical research literature (Salomon et al., 2015), and from novel calculations of expected harms, which employ prior estimates of the utility weights of various health states, of the effects of various medical interventions, and of life expectancy in Nigeria (both absolute and quality adjusted). To create a consistent scale, specific DALY benchmarks are associated with cutoff points that are described in broad clinical terms, capturing the general nature of the harms. Because the DALY consequences of death or disability are higher, and different clinical mistakes are relevant for children, we constructed separate DALY scales for the evaluation of patients who are children (under 18) and patients who are adults.

Physicians first chose one interval of the harm scale, and then on the next screen saw a detailed description of the two scale end-points. They then used a slider to choose the estimated level of harm between those two points. The third-party academic MDs used the same scale for their evaluation. A detailed description is in Appendix C.4.

#### 2.2.3 Health Worker Survey Questions on the LLM Feedback

We asked health workers to give Yes/No answers to three pulse feedback questions about the LLM feedback they got after every patient they saw. In addition, health workers responded to a more general feedback questionnaire when they rotated out of the study. These were intended to elicit whether they perceived LLM assistance as useful and accurate.

### 2.3 Empirical Specification

Our primary analyses obtain the effect of LLM assistance from the within-patient difference between the unassisted and assisted notes. Every outcome is measured for both SOAP notes, and in most cases we estimate

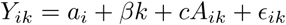

where *i* is the patient and *k* = 1 for the assisted note and zero otherwise, *Y_ik_* is the outcome, and *a_i_* is a patient fixed effect. For outcomes that involve a subjective rating by the physician, *A_ik_* is an indicator for whether SOAP note *k* was shown to the physician first (to control for order effects). We cluster all regressions at the patient level.

## 3 Results

### 3.1 Health worker response to LLM feedback

Health workers made significant changes to their care plans in the assisted SOAP note and generally received LLM feedback positively. In paired comparisons of unassisted and assisted care plans, 33% of pairs differed in the medical test ordered, 41% differed in the clinical indications (diagnoses) recorded, and 54% differed in the prescriptions or drugs recommended (Panel A).

For 95% of patient encounters, the health worker reported that the LLM improved patient documentation and helped provide better patient care (Table 2, Panel B). In our exit surveys, 90-100% of health workers agreed with positive statements about the LLM, such as “I would like to use the LLM in the future” (Panel C). At the same time, 44% of the health workers reported that the LLM responses were too wordy or vague, and 56% agreed that the LLM sometimes made errors. Note that the health care workers rarely reported LLM errors during study encounters (0.5% of encounters), possibly because of the mandatory documentation required to explain reported errors.

**Table 2:**
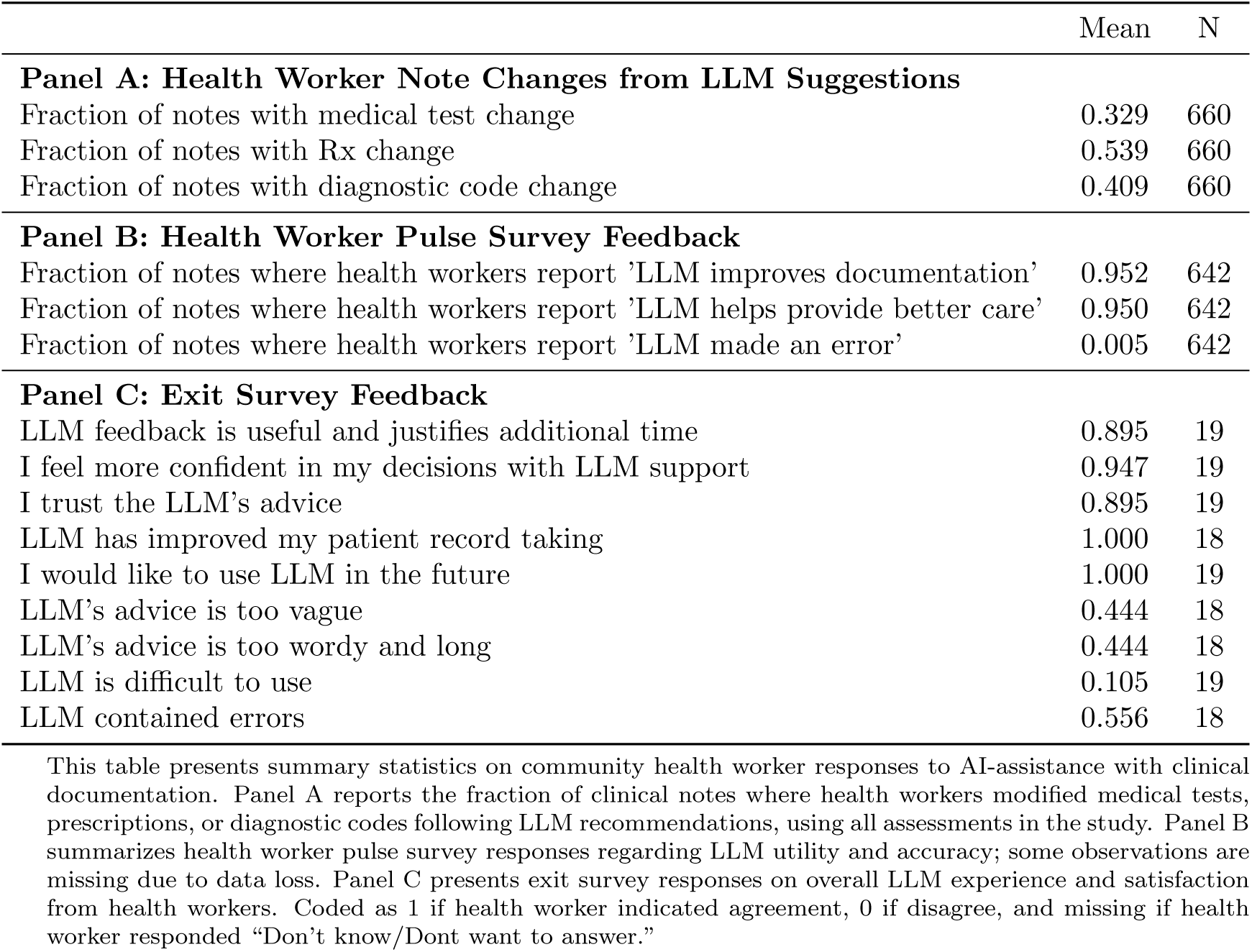
Assessment of the LLM feedback by health workers; pulse survey (after each patient) and exit survey (at the end of the health worker’s study rotation).

|  | Mean | N |
| --- | --- | --- |
| **Panel A: Health Worker Note Changes from LLM Suggestions** |  |  |
| Fraction of notes with medical test change | 0.329 | 660 |
| Fraction of notes with Rx change | 0.539 | 660 |
| Fraction of notes with diagnostic code change | 0.409 | 660 |
| **Panel B: Health Worker Pulse Survey Feedback** |  |  |
| Fraction of notes where health workers report ‘LLM improves documentation’ | 0.952 | 642 |
| Fraction of notes where health workers report ‘LLM helps provide better care’ | 0.950 | 642 |
| Fraction of notes where health workers report ‘LLM made an error’ | 0.005 | 642 |
| **Panel C: Exit Survey Feedback** |  |  |
| LLM feedback is useful and justifies additional time | 0.895 | 19 |
| I feel more confident in my decisions with LLM support | 0.947 | 19 |
| I trust the LLM’s advice | 0.895 | 19 |
| LLM has improved my patient record taking | 1.000 | 18 |
| I would like to use LLM in the future | 1.000 | 19 |
| LLM’s advice is too vague | 0.444 | 18 |
| LLM’s advice is too wordy and long | 0.444 | 18 |
| LLM is difficult to use | 0.105 | 19 |
| LLM contained errors | 0.556 | 18 |

This table presents summary statistics on community health worker responses to AI-assistance with clinical documentation. Panel A reports the fraction of clinical notes where health workers modified medical tests, prescriptions, or diagnostic codes following LLM recommendations, using all assessments in the study. Panel B summarizes health worker pulse survey responses regarding LLM utility and accuracy; some observations are missing due to data loss. Panel C presents exit survey responses on overall LLM experience and satisfaction from health workers. Coded as 1 if health worker indicated agreement, 0 if disagree, and missing if health worker responded “Don’t know/Dont want to answer.”

### 3.2 Physician and MD Evaluations of Unassisted and Assisted Care Plans

Based on the health worker’s responses alone, one might think that the LLM served as a helpful nudge; health workers were able to discern good advice from bad and followed it selectively. However, the evaluation by the physicians who saw the same patients gives a more mixed picture of the value of the LLM feedback.

We begin by analyzing what errors physicians identified in the health worker notes. To do so, we use a set of questions on whether different care plan elements were incorrect.

#### Physician Evaluation of Errors in Care Plan Elements

Table 3 shows the effect of the LLM on identified errors. The outcomes in the first six columns are indicator variables for whether the physician thought that there were missing or incorrect/unnecessary diagnoses, medications, or care instructions. We also code the number of medical tests the health worker ordered that the physician judged to be “unlikely to be clinically useful.”^3^

**Table 3:**
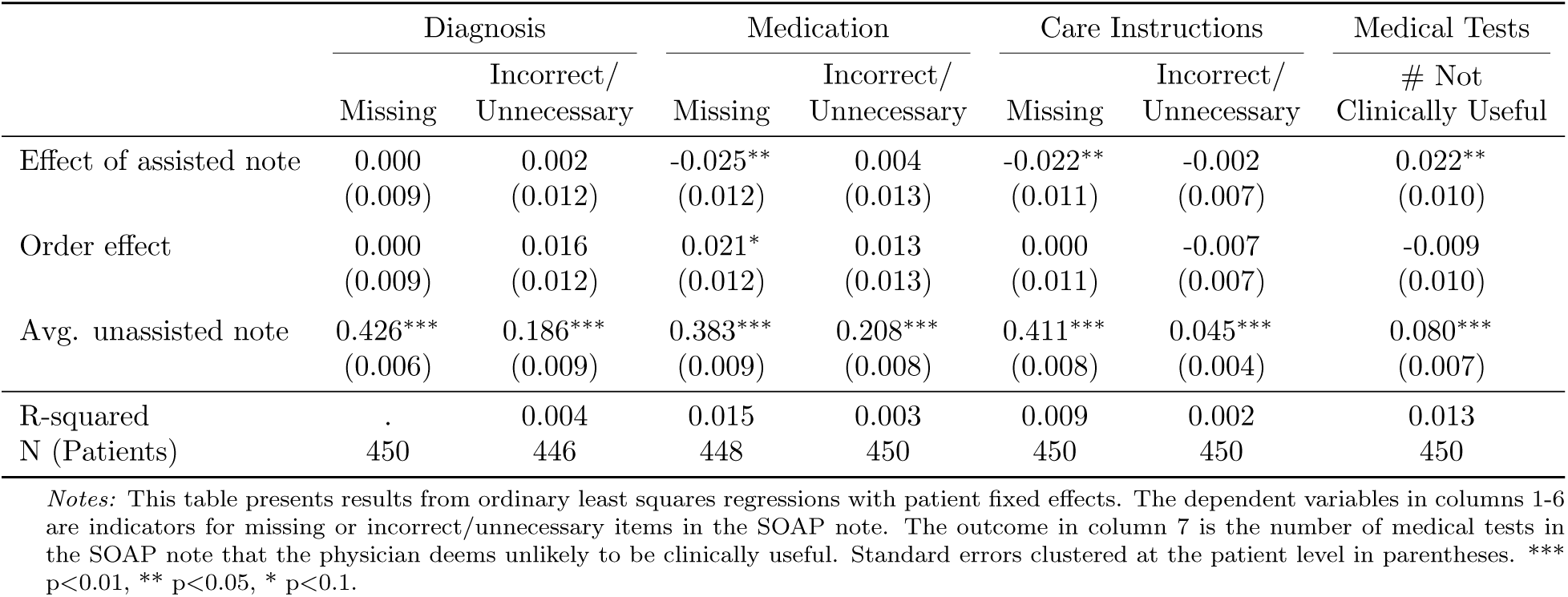
Physician Evaluation of Specific Care Plan Elements

|  | Diagnosis |  | Medication |  | Care Instructions |  | Medical Tests |
| --- | --- | --- | --- | --- | --- | --- | --- |
|  | Missing | Incorrect/<br>Unnecessary | Missing | Incorrect/<br>Unnecessary | Missing | Incorrect/<br>Unnecessary | # Not<br>Clinically Useful |
| Effect of assisted note | 0.000<br>(0.009) | 0.002<br>(0.012) | -0.025**<br>(0.012) | 0.004<br>(0.013) | -0.022**<br>(0.011) | -0.002<br>(0.007) | 0.022**<br>(0.010) |
| Order effect | 0.000<br>(0.009) | 0.016<br>(0.012) | 0.021*<br>(0.012) | 0.013<br>(0.013) | 0.000<br>(0.011) | -0.007<br>(0.007) | -0.009<br>(0.010) |
| Avg. unassisted note | 0.426***<br>(0.006) | 0.186***<br>(0.009) | 0.383***<br>(0.009) | 0.208***<br>(0.008) | 0.411***<br>(0.008) | 0.045***<br>(0.004) | 0.080***<br>(0.007) |
| R-squared | . | 0.004 | 0.015 | 0.003 | 0.009 | 0.002 | 0.013 |
| N (Patients) | 450 | 446 | 448 | 450 | 450 | 450 | 450 |

*Notes:* This table presents results from ordinary least squares regressions with patient fixed effects. The dependent variables in columns 1-6 are indicators for missing or incorrect/unnecessary items in the SOAP note. The outcome in column 7 is the number of medical tests in the SOAP note that the physician deems unlikely to be clinically useful. Standard errors clustered at the patient level in parentheses. \*\*\* p<0.01, \*\* p<0.05, \* p<0.1.

Physicians found many problems with the health workers’ care plans, as shown by the average outcomes for unassisted notes. Approximately 40% of plans are missing key elements like diagnoses that should have been treated, or medications or care instructions patients should have received. In addition to these omissions, 19% of unassisted care plans include incorrect diagnoses, 21% include incorrect or unnecessary medications, and 4% include incorrect or unnecessary care instructions. There are 0.08 medical tests per patient that are not clinically useful.

We find no effect of LLM feedback on diagnoses, incorrect/unnecessary medications or incorrect/unnecessary care instructions, but LLM assistance did reduce the share of notes with missing medications or care instructions by 2.5 percentage points and 2.2 percentage points respectively. These effects are quantitatively small relative to the baseline error rate, but statistically significant. On the other hand, with LLM assistance, the number of tests judged to be not useful increased from 8.0% to 10.2%, a 27.5% increase.

#### Physician Evaluation of Patient Harm

We now turn to whether care plans with LLM-assistance were judged as more or less harmful to patients. Table 4 shows treatment effects of LLM-assistance on various measures of harm, as graded by the physician who evaluated and treated the same patient as the health care worker.

**Table 4:** Physician Assessment of Changes in Quality of Care

|  | Qualitative error/harm |  |  |  | DALY loss |  |  |
| --- | --- | --- | --- | --- | --- | --- | --- |
|  | Any error?* | Short-term harm | Long-term harm | Less harm | Severe loss* | Lower loss* | DALY hours |
| Effect of assisted note | -0.009<br>(0.016) | -0.018<br>(0.017) | -0.009<br>(0.005) | 0.028<br>(0.033) | -0.011*<br>(0.006) | 0.036<br>(0.026) | -147.496<br>(353.375) |
| Order effect | 0.027*<br>(0.016) | 0.018<br>(0.017) | -0.009<br>(0.005) | -0.072**<br>(0.033) | -0.002<br>(0.006) | -0.019<br>(0.026) | 24.001<br>(353.375) |
| Avg. unassisted note | 0.642***<br>(0.012) | 0.524***<br>(0.012) | 0.086***<br>(0.004) | 0.263***<br>(0.023) | 0.054***<br>(0.004) | 0.140***<br>(0.018) | 1017.686***<br>(37.968) |
| R-squared | 0.01 | 0.00 | 0.01 | 0.01 | 0.01 | 0.01 | 0.00 |
| N (Patients) | 450 | 450 | 450 | 448 | 450 | 450 | 450 |

This table presents fixed effects regression estimates of the impact of AI-assisted clinical notes on various measures of medical assessment quality. The dependent variables include binary indicators for treatment errors and harm assessments (columns 1-4), as well as DALY (disability-adjusted life year) loss metrics (columns 5-6). The sample is restricted to assessments evaluated after February 25, when presentation order was randomized to control for order effect. Standard errors clustered at the patient level in parentheses. \* $p < 0.10$ , \*\* $p < 0.05$ , \*\*\* $p < 0.01$ .

The first column outcome is a binary indicator denoting that the care plan was *not* completely appropriate for the patient. This indicator captures all errors regardless of the magnitude of expected harm to the patient. Column 2 captures whether the physician indicated that the health worker’s care plan could cause temporary or short-term harm to the patient, in the form of additional symptoms or missing usual daily activities. The outcome in column 3 is an indicator for whether there was a risk of long-term or permanent harm such as disability or death. Column 4 captures whether the physician judged the care plan (assisted or unassisted) to cause strictly less overall harm than the health worker’s alternative (unassisted or assisted) care plan. This binary outcome was coded as zero in the 51% of cases when the physician judged the care plans as producing equal expected harms. Finally, columns 5 and 6 are constructed from the expected DALY loss estimates the physicians provided for each SOAP note using our two-step harm scale described earlier. Column 5 is an indicator for exceeding the 95th percentile of DALY loss among the unassisted notes, pooling child and adult scales (so the baseline share of the variable is by construction approximately 5%). Column 6 is an indicator for the care plan with the strictly lower DALY loss. This variable differs from the “Less harm” indicator because the DALY scale was collected via a visual scale interface with discrete selectable “notches” so that very small differences in harm could not be recorded. Asterisks denote the pre-specified primary outcomes. While these results are directionally consistent with LLM assistance reducing patient harm, the effect magnitudes for most outcomes are not statistically significant. The “Less harm” and “Lower loss” variables show that in 14-26% of patients, the physician strictly prefers the *unassisted* over the assisted note, and the reverse is only true in a marginally higher share of patients; with many unassisted and assisted notes judged as roughly equivalent. The only meaningful change is the 1.1pp reduction in care plans exceeding the 95th percentile of DALY loss. This effect is significant at the 10% level. These findings suggest that LLM benefits may be concentrated among a small proportion of encounters with the potential for large errors. There appears to be little positive impact for the majority of patients. This result is striking because physicians state that 64% of the unassisted care plans to contain an error, implying ample room for improvement via care plan revisions (Column 1).

Since the estimates suggest sizable order effects in some of the variables, with a harsher judgment on the note randomly chosen to be presented first (SOAP A), we repeat the estimates including only SOAP A and comparing assisted and unassisted notes in the cross section, shown in Appendix Table B.1. The results vary somewhat across outcomes but do not fundamentally alter the findings in terms of the magnitude of quality of care improvements by the LLM. We also repeated this analysis with short-term and long-term harm assessed by another LLM (GPT-5) rather than by the treating physician; findings are similar, with small but insignificant harm reduction, albeit a much higher baseline rate of harm and no order effects (Appendix Table B.2). Finally, noting that physicians are somewhat better than chance at identifying the assisted SOAP note after seeing both notes (Appendix Table B.3), we assess whether issues with incomplete blinding could have caused bias in our results, for example because physicians oppose the use of LLMs in patient care. Physicians tended to rate a care plan more favorably when they believed the plan to be LLM-assisted. However, the estimated effects of LLM assistance remained nearly unchanged when regression models were modified to include a binary variable denoting the physician’s guess regarding which note was LLM-assisted (Table B.4 in the Appendix).

#### MD Evaluation of Harm

To supplement the treating physicians’ ratings, third-party MDs re-rated care plans for 59 patients in double review. For each study encounter, two MDs were initially presented with physician notes and all medical test results. MDs were then asked whether the physician made any serious errors; they reported such errors for only 2 encounters. They then evaluated the health worker notes in random order, in parallel fashion to the evaluations by the treating physician. The 59 patients were selected for MD review based on either a high loss rating from the physician in the unassisted note or a strong increase or decline in the loss rating. Our main interest is not generalizing from this (selected) subsample to the full sample, but understanding how MDs and physicians compare in their evaluations within this pivotal subsample. Table B.5 in the Appendix documents that there is substantial disagreement between the physicians and MDs on the presence of short- and long-term harm as well as preference between the two notes.

Table 5 shows treatment effects for the MD ratings in Panel A and the physician ratings in the same sample of patients in Panel B. Because two MDs rated each note, standard errors were clustered at the patient level. However, results are very similar if each MD ratings are analyzed separately (not shown). Physician ratings in Panel B confirm this subsample has much larger baseline rates of error and harm than the full sample, as well as larger – but still statistically insignificant – estimated effects of LLM-assistance. By comparison, the MD ratings indicate a much stronger preference for the assisted SOAP note, with significant reductions in error and harm rates, and a much higher share of assisted notes rated as causing “Less harm” and “Lower loss” than the unassisted note. A manual review suggests that the differences are driven by patients with uncontrolled hypertension. The LLM seems to change the health worker’s care plan for hypertensive patients, but the MDs seem to ascribe greater health gains than the physicians to the additional care that the LLM recommends. This divergence between the MD and treating physician’s assessment might reflect superior knowledge about future risks on the part of more highly trained MDs. Alternatively, the treating physicians might ascribe lower health returns to the LLM-induced care plan updates based on direct interactions with the patient that inform expectations regarding the long-term success of more aggressive treatment.

**Table 5:** MD and Physician Assessment of Changes in Quality of Care in the Same Sample

|  | Qualitative error/harm |  |  |  | DALY loss |  |
| --- | --- | --- | --- | --- | --- | --- |
|  | Any error?* | Short-term harm | Long-term harm | Less harm | Severe loss* | Lower loss* |
| **Panel A: MD ratings** |  |  |  |  |  |  |
| Effect of assisted note | -0.158***<br>(0.044) | -0.125***<br>(0.044) | -0.084***<br>(0.026) | 0.254***<br>(0.066) | -0.007<br>(0.014) | 0.135***<br>(0.043) |
| Order effect | -0.068<br>(0.044) | -0.036<br>(0.044) | -0.013<br>(0.026) | 0.058<br>(0.066) | -0.025*<br>(0.014) | 0.010<br>(0.043) |
| Avg. unassisted note | 0.634***<br>(0.033) | 0.500***<br>(0.033) | 0.201***<br>(0.019) | 0.126***<br>(0.045) | 0.063***<br>(0.011) | 0.292***<br>(0.034) |
| R-squared | 0.12 | 0.07 | 0.09 | 0.12 | 0.03 | 0.08 |
| N (MD-patient pair) | 118 | 118 | 118 | 117 | 118 | 118 |
| **Panel B: Physician ratings (MD sample)** |  |  |  |  |  |  |
| Effect of assisted note | -0.036<br>(0.048) | -0.050<br>(0.062) | -0.065<br>(0.040) | -0.071<br>(0.119) | -0.016<br>(0.016) | -0.005<br>(0.116) |
| Order effect | 0.036<br>(0.048) | -0.014<br>(0.062) | -0.065<br>(0.040) | 0.071<br>(0.119) | -0.016<br>(0.016) | 0.102<br>(0.116) |
| Avg. unassisted note | 0.932***<br>(0.033) | 0.905***<br>(0.040) | 0.471***<br>(0.031) | 0.407***<br>(0.084) | 0.160***<br>(0.016) | 0.341***<br>(0.082) |
| R-squared | 0.02 | 0.01 | 0.09 | 0.01 | 0.03 | 0.01 |
| N (Patients) | 59 | 59 | 59 | 59 | 59 | 59 |

This table presents fixed effects regression results comparing academic MD and on-site physician assessments of care quality for the same sample of 59 patients. These patients were selected for MD review based on either a high DALY loss rating from the physician in the unassisted note or a substantial change in the physician’s loss rating between unassisted and assisted notes. Each patient was independently reviewed by two MDs and one physician, with each reviewer evaluating both the unassisted and assisted SOAP notes. Panel A shows MD ratings across six outcome measures: qualitative errors/harms (any error, short-term harm, long-term harm) and DALY loss outcomes (less harm, severe loss, lower loss), based on 118 MD-patient pairs (59 patients × 2 MDs per patient). Panel B presents physician ratings for the same 59 patients. The regressions use reviewer-patient pair fixed effects to compare assisted versus unassisted notes within the same patient. The reported N refers to the number of reviewer-patient pairs. Standard errors are clustered at the patient-reviewer level for MD ratings (Panel A) and clustered at the patient level for physician ratings (Panel B). Standard errors are reported in parentheses. \* $p < 0.10$ , \*\* $p < 0.05$ , \*\*\* $p < 0.01$ .

### 3.3 Testing and Treatment Misallocation

A key measure of quality is whether the LLM improved the allocation of medical care, in the sense that appropriate treatments for a given condition are matched to patients with that specific condition. Such efficient allocation usually requires correct identification of a patient’s condition, i.e. diagnosis. We use the results from our expanded testing for malaria, UTI, and anemia to determine whether the LLM improved the allocation of medical testing and, ultimately, treatment, for these conditions.

#### Effects on Test Detection Rates

Panel A of Table 6 shows the effect of the LLM on changes in laboratory tests that turned out negative vs. those that turned out positive for the three conditions. The variable “Tested Negative” is defined as the health worker ordering a test for a patient with no symptoms or a negative test result, and “Missed Positive” is defined as not ordering a test for a patient with a positive test result.^4^ Panel B also shows average “tested negatives” and “missed positives” and the absolute testing rates in the health worker and physician care plans, along with the total share of patients with symptoms and an abnormal test for each condition, i.e. the assumed share of patients who should have received treatment in this sample.^5^

**Table 6:**
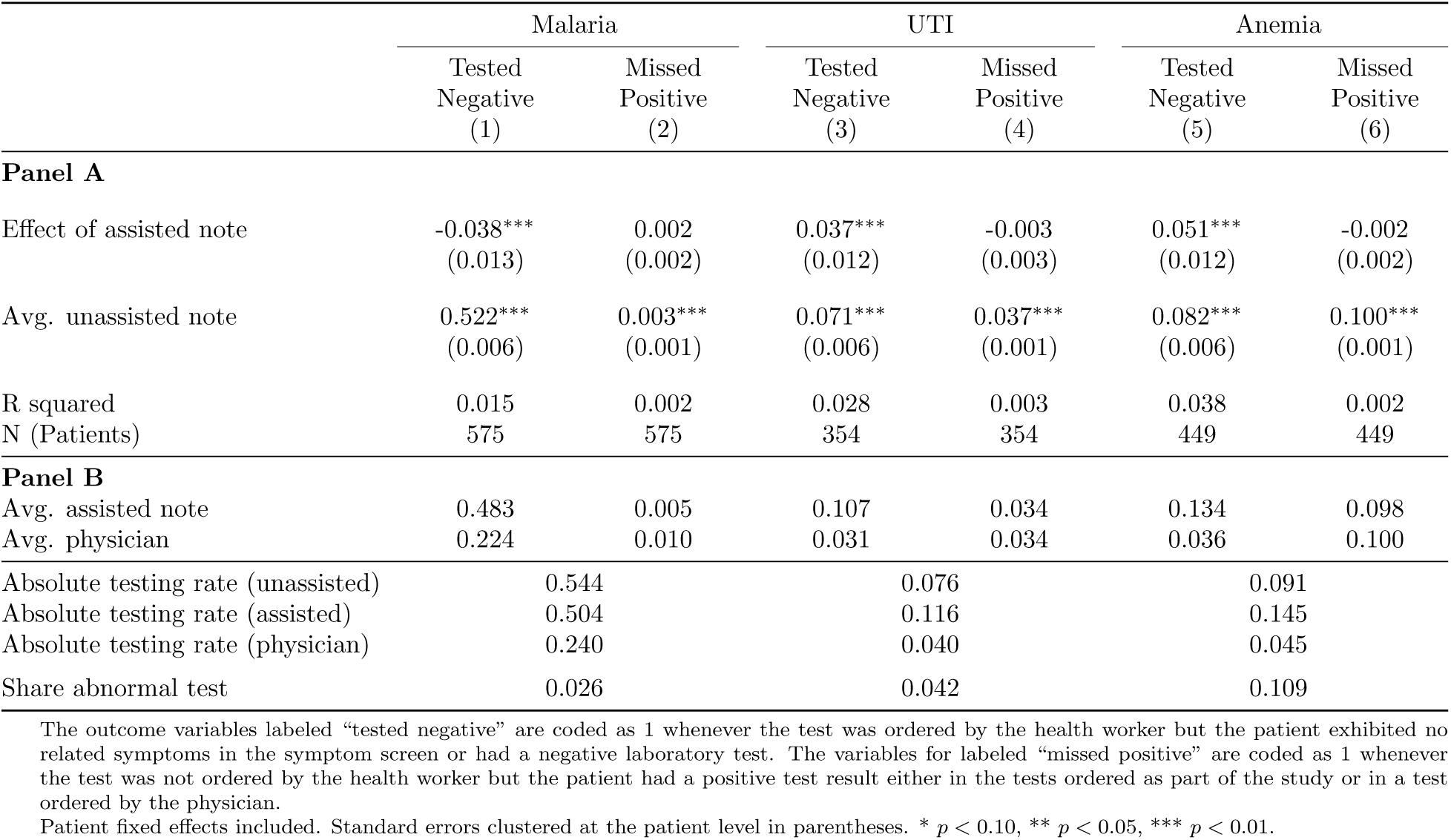
Tested Negatives and Missed Positives for Malaria, UTI and Anemia.

A first observation is that physicians are generally more efficient in the use of medical tests than health workers: despite lower absolute testing rates for all conditions and especially for malaria, their “missed positive” rates are almost identical (and consequently, their “tested negative” rates are significantly lower). The average of the unassisted note shows that health workers order a test that ends up negative in 32.7% of patients for malaria, where only 2.6% of patients have positive tests.^6^ There are very few “missed positives” for malaria. Conversely, while 4.2% of urine test results are consistent with UTI and 10.9% of blood tests are consistent with anemia, both health workers and physicians fail to order tests for most of these cases. While the LLM leads to the addition of a significant number of tests – 4% of patients for UTI, and 5.4% for anemia – almost all the additional tests turned out negative, suggesting that the LLM does not identify the right patients for testing.

We conduct a variety of robustness checks. First, a share of patients who were eligible for medical tests based on their symptoms did not consent to the test and had to be excluded from the analysis. This may lead us to overestimate the share of tested negatives and underestimate the missed positives, given that these patients exhibited symptoms. In Appendix Table B.7 we therefore restrict the sample to patients who were actually tested (i.e. exhibited symptoms and qualified for our screening tests, or were tested by the physician) as a proxy bound on effect sizes. The effects are qualitatively similar and there is no significant reduction in missed positives in this subsample.

Second, negative test results are part of an efficient testing strategy and are not sufficient evidence to indicate poor decision-making. To address this issue, we define alternative outcome variables that use the physician’s decision to test as a benchmark (see Appendix Table B.8). The results are remarkably similar: both the LLM-induced increase in anemia and UTI tests and the reduction in malaria testing are in patients that the physicians did not test, and so tested negatives relative to the physician is increased for the former but reduced for the latter (benchmarking to the physician, we do detect a statistically significant reduction in missed positives for anemia).

Finally, since the smaller post-February 25 sample has slightly different rates in serious illness (see 2), we replicate the analysis in the smaller sample and confirm that the results remain qualitatively unchanged (Appendix Table B.9).

#### Cost-Benefit and Diagnostic Yield

Another way to address testing efficiency is to ask whether LLM-induced changes to test utilization pass a cost-benefit analysis: an increase in testing for UTI and anemia that identifies even a small number of additional positive cases could be worthwhile if tests are cheap relative to the value of identifying positive patients. Conversely, identifying one fewer malaria-positive patient could be beneficial if it saves many negative tests. In general, the low-cost of testing relative to the harms from missed positives precludes drawing any firm conclusions about costs and benefits.^7^

For malaria, LLm assistance removed 30 negative tests and one positive test. Strikingly, the removal of the positive malaria test occurred as a result of user error. Although the LLM had suggested that the CHEW “ensure the malaria RDT test is positive” before initiating treatment, the malaria test was instead removed from the care plan. The resulting care plan still included antimalarial medication.

For anemia, the assisted note added 27 anemia tests, of which one was positive (see Appendix Table B.6). The test yield among marginally tested patients is 3.7%, below the 11% we see in our broad screening test, suggesting that the LLM had poor diagnostic accuracy.

For UTI, the assisted note added 15 UTI tests, of which one was positive. This yield of 6.7% among the marginally tested patients is slightly above the 4% we see in our screening test. Given the low cost of the tests (50 cents), these additional tests could be desirable from a cost-benefit perspective even though they were mostly negative. However, a lower valuation of health gains, or cost estimates that account for patient time or community antibiotic resistance may tip the cost-benefit balance against the additional testing.

#### Misallocation of Treatment

Ultimately, what matters for quality of care is whether patients receive the right treatment. Misallocation of treatment is a persistent problem in many low-resource settings (e.g. (Das et al., 2016; Sulis et al., 2020; Wagner et al., 2024)). Undertreatment may occur if a given condition is not detected or if the provider gives the wrong drug. Overtreatment for a condition may occur if the provider gives presumptive treatment without a test, or treats despite a negative test result.^8^ Overtreatment can simply be a result of misdiagnosis, but may also reflect patient demand or provider incentives to up-sell (Currie et al., 2014; Lopez et al., 2022).

While we cannot identify misallocation of treatment for all diseases and all care plans, we can ask whether treatment for malaria, UTI and anemia was correctly received (or not received) in our screened sample. As illustrated in Table 7, there were no statistically significant reductions in treatment misallocation associated with LLM assistance. Our results on medical testing are in line with undertreatment rates remaining unchanged (note that the study period did not overlap with malaria season, which may have removed a potential cause of undertreatment among the conditions we screened). It is also worth keeping mind that we may underestimate the incidence of overtreatment in our setting, because health workers knew they were under scrutiny and at the same time were not actually making decisions about patient care, so they were not subject to patient pressure or a motivation to increase clinic revenue. Accordingly, most health workers created conditional treatment plans that indicated they would only give treatment with a confirmatory test result. Our results partly confirm this intuition. Overall we find moderate baseline rates of misallocation and the LLM feedback does not improve these rates significantly. The results are similar in the smaller, fully randomized sample (see Appendix Table B.10).

**Table 7:** Treatment Misallocation for Malaria, UTI and Anemia.

|  | Overall* | Malaria | UTI | Anemia |
| --- | --- | --- | --- | --- |
| Effect of assisted note | -0.004<br>(0.004) | -0.002<br>(0.006) | -0.006<br>(0.004) | -0.007<br>(0.007) |
| Avg. unassisted note | 0.074***<br>(0.002) | 0.030***<br>(0.003) | 0.060***<br>(0.002) | 0.144***<br>(0.004) |
| R squared | 0.001 | 0.000 | 0.006 | 0.002 |
| N (Patients) | 1,416 | 604 | 358 | 454 |

Misallocation is coded as 1 when there is an intent to treat not conditioned on a positive test but the test is negative; or when the incorrect drug or dosage was prescribed or no treatment was prescribed and the test is positive. When the test result is missing, misallocation is coded as 1 when there is an unconditional incorrect drug or dosage, and 0 when a conditional plan specifies correct drug and dosage and otherwise as missing. Overall misallocation is prespecified in the pre-registration plan and at the patient-test level. Patient fixed effects included. Standard errors clustered at the patient level in parentheses. \* $p < 0.10$ , \*\* $p < 0.05$ , \*\*\* $p < 0.01$ .

### 3.4 Concordance Between Health Worker and Physician Diagnosis and Treatment

Yet another approach to assessing quality of care is to assess whether LLMs make the care plans of health workers more like those of treating physicians. We already assessed subjective alignment in Table 3, filtered through the physician’s assessment of missing vs. unnecessary care elements. Assessing objective alignment amounts to assuming that any increased alignment between health workers and physicians is suggestive of LLM benefits. Given physicians’ longer training and broader scope of practice, it is reasonable to believe that physician care plans are higher-quality care than those of community health workers, although this approach cannot weigh the importance of each care plan element.

In order to assess objective matches in diagnosis and treatment elements, we constructed categories that grouped similar diagnostic codes and prescriptions. We then created variables measuring discordance between physician and health worker care plans with respect to these grouped elements. These discordance measures are defined as the count of items in the health worker note that were not included in the physician note (overprescription/overdiagnosis) and the number of items in the physician note not included in the health worker note (underprescription/underdiagnosis).^9^

In Table 8 we report indicators for any overprescription and underprescription and any overdiagnosis and underdiagnosis to facilitate comparison with the subjective assessments of missing vs. unnecessary items by the physician in Table 3. The results show high rates of mismatch and small improvements across the different indicators, with only the effect on any underprescription marginally significant. In Appendix Table B.11 we also report effects on counts of mismatched elements, where we see marginally significant evidence of *increased* overprescribing, suggesting that if the LLM overprescribes, it is more likely to recommend several treatments. When replicating Table 8 in the smaller sample, the results remain unchanged (Appendix Table B.12).

**Table 8:**
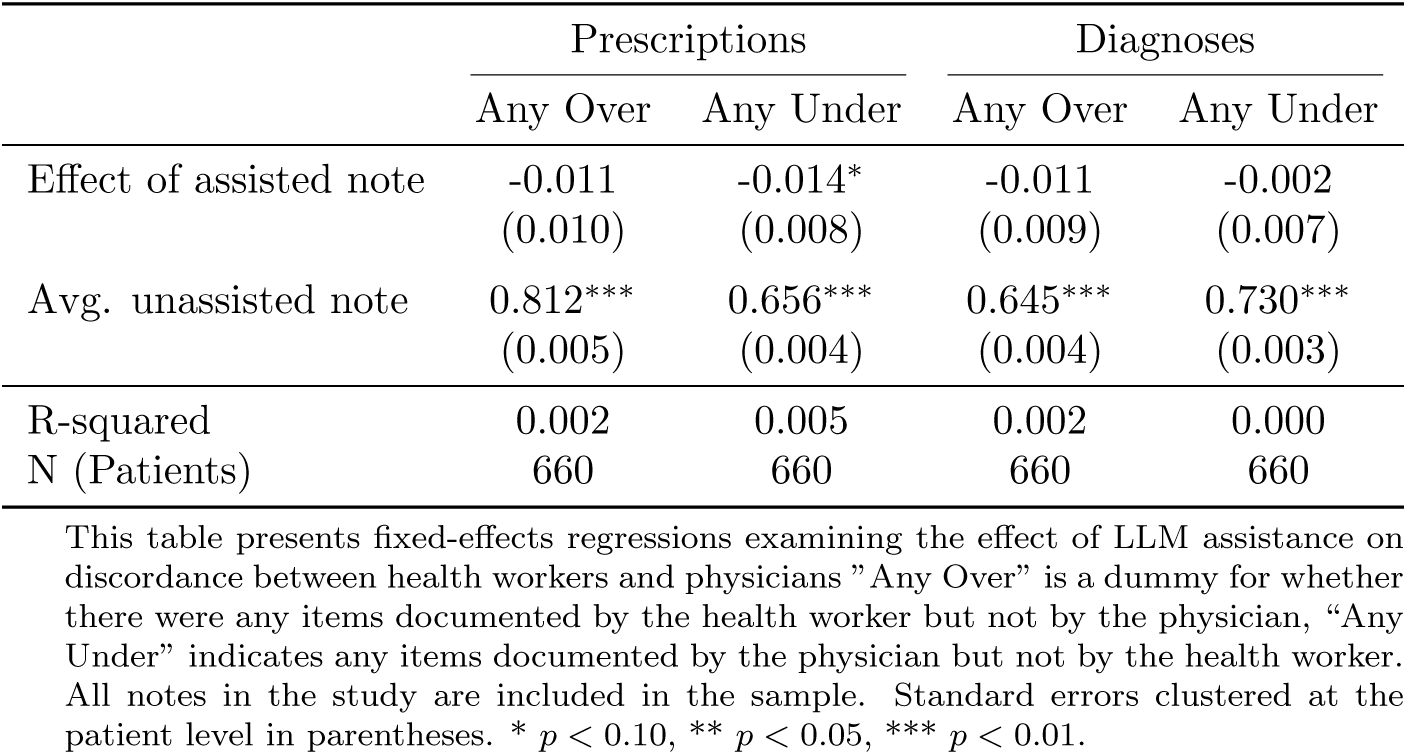
Health Worker-Physician Objective Discordance

|  | Prescriptions |  | Diagnoses |  |
| --- | --- | --- | --- | --- |
|  | Any Over | Any Under | Any Over | Any Under |
| Effect of assisted note | -0.011<br>(0.010) | -0.014*<br>(0.008) | -0.011<br>(0.009) | -0.002<br>(0.007) |
| Avg. unassisted note | 0.812***<br>(0.005) | 0.656***<br>(0.004) | 0.645***<br>(0.004) | 0.730***<br>(0.003) |
| R-squared | 0.002 | 0.005 | 0.002 | 0.000 |
| N (Patients) | 660 | 660 | 660 | 660 |

This table presents fixed-effects regressions examining the effect of LLM assistance on discordance between health workers and physicians. "Any Over" is a dummy for whether there were any items documented by the health worker but not by the physician, "Any Under" indicates any items documented by the physician but not by the health worker. All notes in the study are included in the sample. Standard errors clustered at the patient level in parentheses. \* $p < 0.10$ , \*\* $p < 0.05$ , \*\*\* $p < 0.01$ .

## 4 Mechanisms

Overall, our results paint a mixed picture. Both the objective results on concordance and the physician evaluations suggest at best small benefits of LLM assistance, and substantial variability.

Why did the LLM feedback not move the health worker’s care plans closer to that of the treating physician? In this section, we directly analyze the LLM feedback health workers received, as well as the role of health worker discretion in adopting LLM recommendations and the role of incomplete data elicitation from patients by health workers.

We first check whether the LLM provided actionable recommendations that *could* have led to more concordance with the physician. We ask GPT-5 to identify the specific recommendations the LLM made for the health worker, label them by whether the LLM suggests changing a part of the care plan (e.g., “Consider an h. pylori test”) or keeping it the same (e.g., “Continue the antibiotic treatment plan”), and classify which part of the treatment plan they refer to: prescriptions, medical (lab) tests, or advice.

Next, we ask whether the LLM recommendation would make the health worker note *more similar* to that of the physician – meaning whether following it would completely or partially reduce any differences between health worker and physician (e.g., add a missing physician item, remove an incorrect health worker item) – or *less similar* – e.g., introduce a new test/drug not in the physician note, or alter a shared correct element to diverge.

The top panel of Table 9 shows the average counts of recommendations per patient that included some change, broken down by whether those changes encourage a note to be more or less similar to the physician. The LLM makes many recommendations – 3.8 per patient – despite the prompt instructions to be concise and focus on the most important issues. 53% of these recommendations would make the health worker note more similar to the physician. Another 35% would make the notes less similar and 12% of the recommendations do neither; these might suggest for example to replace a medication not in the physician note with another one not in the physician note.

**Table 9:**
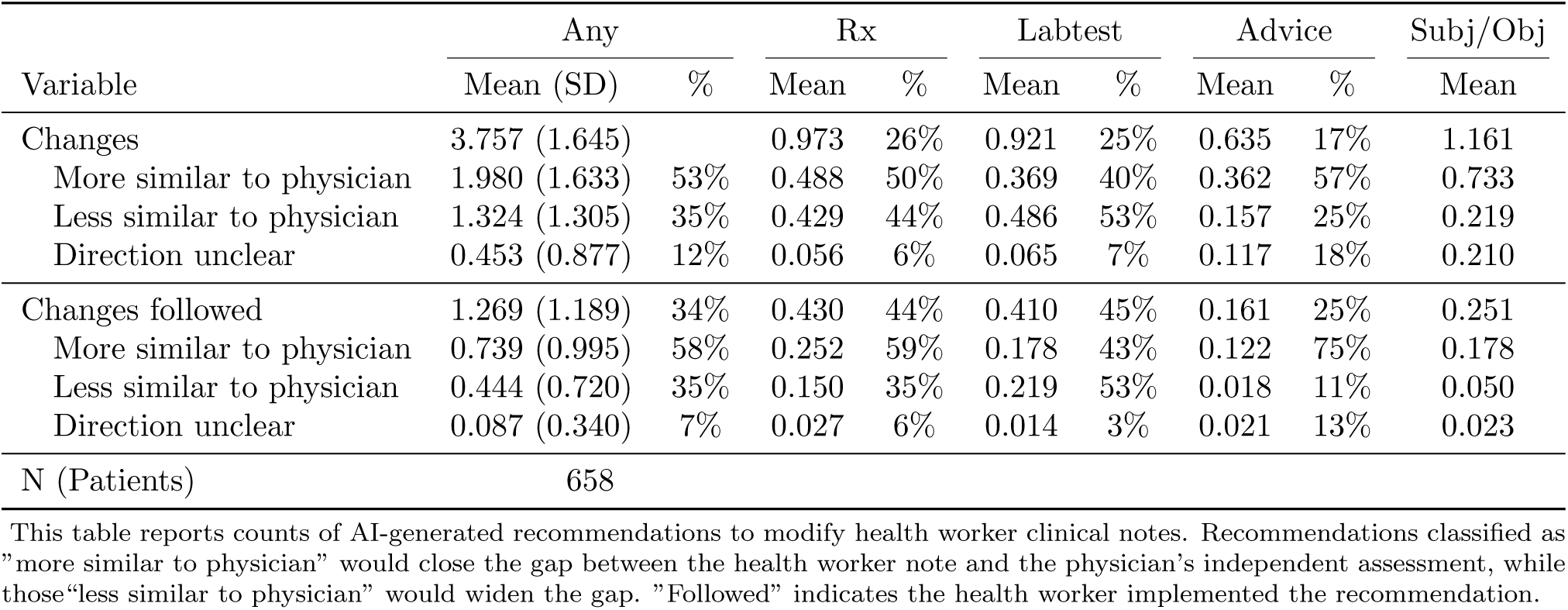
LLM Recommendations: Alignment with Physician and Health Worker Follow-Through

We next conduct two pieces of analysis to understand the role of the health worker better. First, we let GPT-5 label each recommendation as “followed” or “ignored” based on the changes from the unassisted to the assisted health worker note.^10^ Table 9 shows that the health workers only followed 34% of the LLM’s recommendations. They were on average slightly more likely to follow good recommendations than bad: 58% of the followed changes increased concordance with the physician, while 42% had an unclear effect or decreased concordance. The share of (followed) changes that made the notes “more similar” varied across categories from 43% for lab tests to 75% for advice. The health workers were in all categories slightly better than chance in identifying and following the “more similar” rather than the “less similar” and “unclear” recommendations, but the “human in the loop” is far from a perfect safeguard against the 47% of LLM recommendations that are unhelpful (measured against the physician as the benchmark). The health workers had the best ratio of more to less similar followed recommendations in the “Advice” category; on-site physicians in their subjective reviews may have weighted good advice less heavily than appropriate prescriptions or treatment.

Second, we want to test the hypothesis that the unassisted SOAP note does not contain sufficient relevant patient information for the LLM to provide useful feedback (e.g., by flagging inconsistencies or unaddressed signs and symptoms). To do so we construct a synthetic SOAP note composed of the Assessment and Plan from the health worker note, but the Subjective and Objective components of the physician note. In other words, we combine the physician’s description of the patient’s appearance, signs, and symptoms with the health worker’s lab tests ordered, prescriptions and advice, and diagnosis. We then regenerate the LLM feedback with the original prompt the LLM used in the experiment and repeat the first part of the exercise above to measure the extent to which LLM recommendations based on the *synthetic note* would make the health worker’s care plan more similar to the treating physician’s. If the physician records more detailed information than the health worker, it might enable the LLM to identify inconsistencies in the health worker’s care plan and improve the alignment of the LLM’s recommendations with the physician. To test this, we run a fixed effect regression with the original and the synthetic SOAP notes where the outcome *Y_ik_* is the number of LLM recommendations in a given category (“more similar” or “less similar”, by component) and the independent variable *T_k_* is an indicator for the synthetic note.

Table 10 shows the change in the number of recommendations, in total and split into “more similar” and “less similar.” We see a decrease of “less similar” recommendations with the synthetic note by -0.078 and a small shift from “less” to “more” similar advice in the Rx category (both not statistically significant). The synthetic note significantly shifts recommendations on lab tests from the “less similar” to the “more similar” category. This is suggestive evidence that the subjective and objective information the health worker records partly contributes to leading the LLM “down the wrong path.” However, even with the treating physician’s S-O portion of the note, the LLM continues to make on average more than one recommendation per patient that would move the health worker and physician further apart. We cannot distinguish whether this is because the physician’s S-O records on the patient are also incomplete, or because the LLM interpretation of the S-O contents is different from the physicians.

**Table 10:** Synthetic Note Analysis: Effect of Including Physician’s Patient Observations on Quality of LLM Feedback.

|  | *Any Change* |  |  |  |  |
| --- | --- | --- | --- | --- | --- |
|  | Any | Rx | Lab Test | Advice | Rx+Lab |
| Effect of Physician’s S-O | 0.124<br>(0.089) | -0.012<br>(0.045) | 0.031<br>(0.037) | 0.024<br>(0.044) | 0.018<br>(0.058) |
| Baseline (Health Worker’s S-O) | 3.756***<br>(0.064) | 0.973***<br>(0.034) | 0.921***<br>(0.029) | 0.635***<br>(0.033) | 1.894***<br>(0.044) |
|  | *More Similar to Physician* |  |  |  |  |
|  | Any | Rx | Lab Test | Advice | Rx+Lab |
| Effect of Physician’s S-O | 0.007<br>(0.081) | 0.030<br>(0.037) | 0.096***<br>(0.030) | -0.009<br>(0.034) | 0.126**<br>(0.051) |
| Baseline (Health Worker’s S-O) | 1.980***<br>(0.064) | 0.488***<br>(0.028) | 0.369***<br>(0.023) | 0.362***<br>(0.027) | 0.857***<br>(0.039) |
|  | *Less Similar to Physician* |  |  |  |  |
|  | Any | Rx | Lab Test | Advice | Rx+Lab |
| Effect of Physician’s S-O | -0.078<br>(0.062) | -0.020<br>(0.033) | -0.068**<br>(0.030) | 0.006<br>(0.023) | -0.088*<br>(0.045) |
| Baseline (Health Worker’s S-O) | 1.324***<br>(0.051) | 0.429***<br>(0.025) | 0.486***<br>(0.024) | 0.156***<br>(0.017) | 0.915***<br>(0.036) |
| N (Patients) | 659 | 659 | 659 | 659 | 659 |

This table reports the impact of providing the physician’s subjective-objective (S-O) information to the LLM compared to providing only the health worker’s S-O information, estimated using patient fixed effects regressions. Panel A shows the total number of recommendation changes by category. Panel B presents the number of recommendation that made the LLM’s assessment more similar to the physician’s final assessment. Panel C shows the number of recommendation that made the LLM’s assessment less similar to the physician’s final assessment. Each column represents different types of recommendations: any change, prescriptions (Rx), lab tests, advice, and the sum of Rx and lab test changes. Standard errors are clustered at the patient level. Statistical significance: \* $p < 0.10$ , \*\* $p < 0.05$ , \*\*\* $p < 0.01$ .

## 5 Conclusion

We study the use of LLMs for real-time clinical decision support in a low resource setting. We find that health workers value the LLM feedback and respond with many changes to their care plans, and in a subsample of patients, care quality seems to improve according to MDs who review a record of the patient interaction. Our main contribution is an analysis of care plan reviews by physicians who evaluated and treated the patient during the same clinic encounter, as well as medical test results for three commonly treated health conditions.

These new methods of measurement suggest a more mixed picture. Objective testing suggests that LLMs reduce tested negatives for malaria but increase tested negatives for anemia and UTIs while identifying few undetected abnormal cases of any condition. The treating physicians also mostly do not judge the LLM-assisted care plans as improved, and objective measures of concordance between health worker and physician show almost no change. The discrepancy between the treating physicians and reviewing MDs may reflect differences in expertise, but could also be due to better information or a different assessment of the health consequences of the changed care plan on the part of the treating physicians. In the subsample reviewed by the MDs, a major benefit of the assisted notes was better identification of hypertensive patients; the on-site physicians might believe that addressing the elevated blood pressure did not contribute to solving the patient’s acute problem, was not a patient priority, or would not be effective.

Exploring mechanisms, we find that LLMs make many recommendations for changes that do not move the health worker’s care plan closer to the physician, and while the health workers follow only about one third of the LLM recommendations, they are only slightly more likely to follow advice that improves alignment with physicians. There is suggestive evidence that some of the problem can be attributed to insufficient patient information recorded by the health workers, but even when supplying the same, signs and symptoms the treating physicians recorded, the LLM still makes many recommendations that would make the care provided by the health workers less similar to that provided by the physician.

There are two main takeaways from our results, one practical and one methodological. Current generation LLMs seem capable of modestly improving some aspects of healthcare delivery, although the benefits are mixed when assessed using objective tests or by physicians who evaluated and treated the patient. While we do not observe evidence that LLMs harm patient care, the absence of consistently measured and large beneficial effects suggests that deployment of LLMs in clinical decision support might not (yet) be an urgent policy priority.

Methodologically, our results highlight the importance of assessing care through multiple means. Comparison with medical guidelines or third-party retrospective review may deliver a very different picture from an evaluation based on objective test results or comparison with the care decisions by physicians who see and treat the same patient.

## Data Availability

All data produced in the present study are available upon reasonable request to the authors.

## Supplemental Materials

### A LLM prompt

The following is the prompt used to request LLM feedback. A CHEW SOAP note extracted from the EMR is appended to this prompt and sent to the LLM:

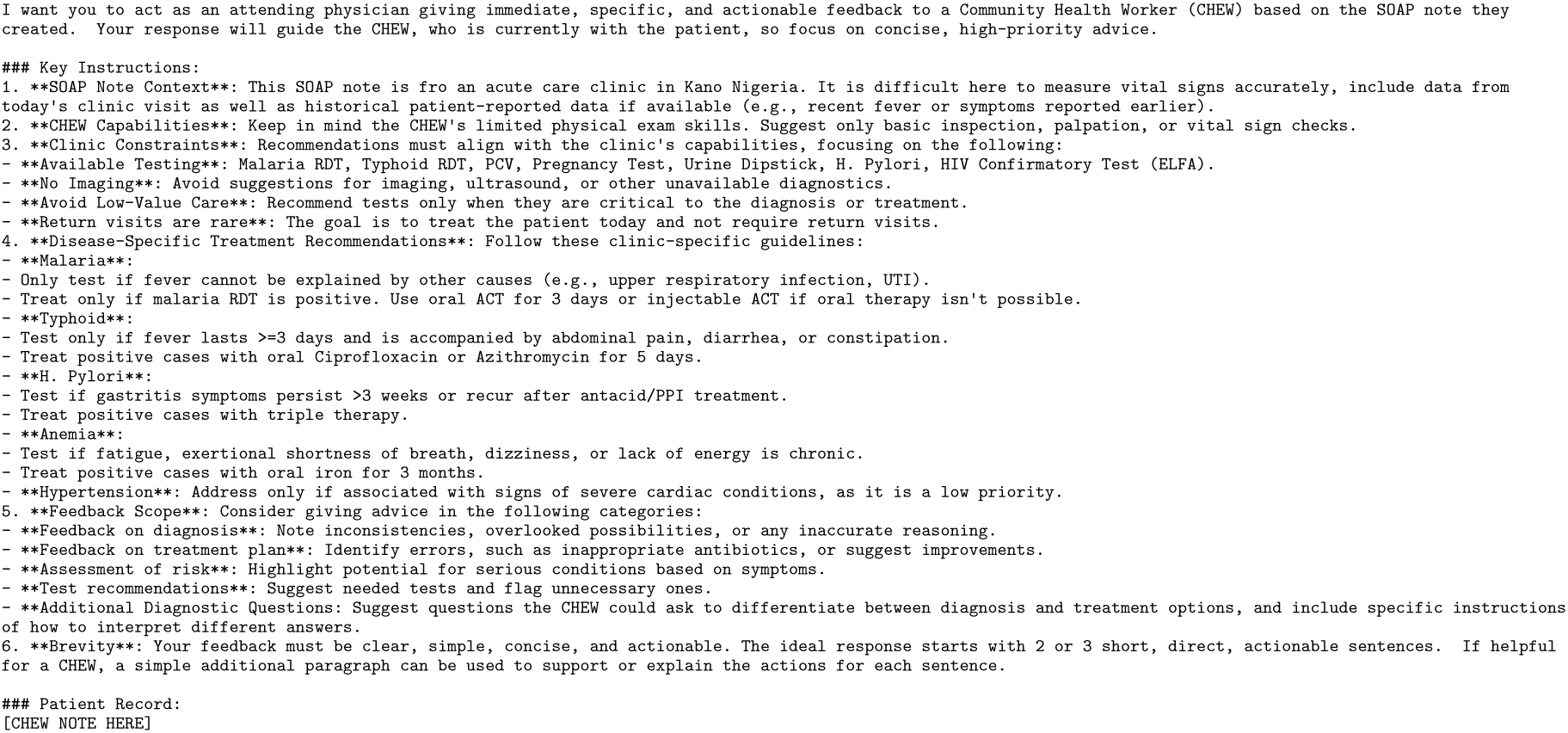

The LLM response is received and forwarded to the CHEW in real time. An example LLM response is:

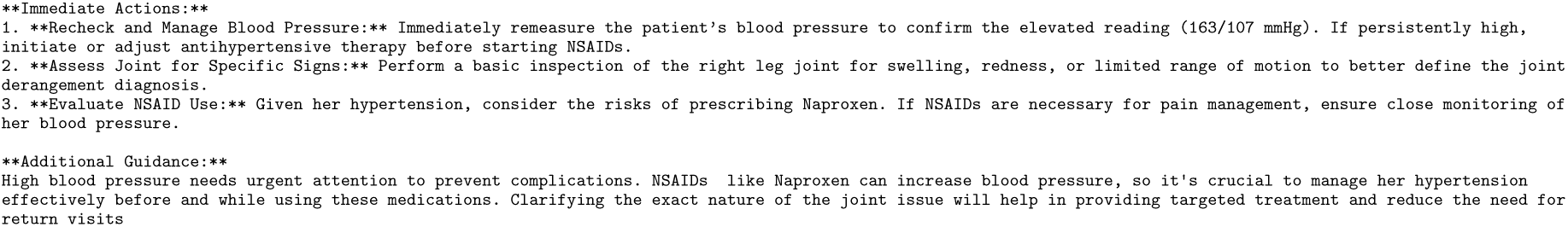

### B Additional Figures and Tables

**Table B.1:** Physician Assessment of Quality of Care, SOAP Note A only.

|  | Qualitative error/harm |  |  |  | DALY loss |  |
| --- | --- | --- | --- | --- | --- | --- |
|  | Any error?* | Short-term harm | Long-term harm | Less harm | Severe loss* | Lower loss* |
| Effect of assisted note | -0.016<br>(0.045) | 0.039<br>(0.047) | -0.043*<br>(0.025) | 0.018<br>(0.038) | -0.024<br>(0.020) | 0.052<br>(0.033) |
| Avg. unassisted note | 0.673***<br>(0.032) | 0.514***<br>(0.034) | 0.095***<br>(0.020) | 0.196***<br>(0.027) | 0.059***<br>(0.016) | 0.114***<br>(0.021) |
| R-squared | 0.00 | 0.00 | 0.01 | 0.00 | 0.00 | 0.01 |
| N (cases) | 450 | 450 | 450 | 448 | 450 | 450 |

This table presents OLS regression estimates of the impact of AI-assisted clinical notes on alternative measures of treatment quality. The dependent variables include binary indicators for treatment errors and harm assessments (columns 1-4), as well as DALY (disability-adjusted life year) loss metrics (columns 5-6). The sample includes only the first assessment (SOAP A) for each patient-condition pair and is restricted to assessments evaluated after February 25, when presentation order was randomized to control for order effects. Robust standard errors in parentheses. \* $p < 0.10$ , \*\* $p < 0.05$ , \*\*\* $p < 0.01$ .

**Table B.2:** LLM Assessment of Changes in the Quality of Care

|  | Qualitative error/harm |  |
| --- | --- | --- |
|  | Short-term harm | Long-term harm |
| Effect of assisted note | -0.011<br>(0.011) | -0.020<br>(0.026) |
| Order effect | 0.007<br>(0.011) | 0.020<br>(0.026) |
| Avg. unassisted note | 0.959***<br>(0.007) | 0.572***<br>(0.018) |
| R-squared | 0.00 | 0.00 |
| N (Patients) | 450 | 376 |

This table presents fixed effects regression estimates of the impact of AI-assisted clinical notes on medical officer assessments of potential harm from treatment errors. The dependent variables are binary indicators for whether the treatment error could cause short-term harm (column 1) or increase the risk of long-term harm (column 2). The sample is restricted to assessments evaluated after February 25, when presentation order was randomized to control for order effects. "Unclear" responses for long-term harm are coded as missing. Standard errors clustered at the patient level in parentheses. \* $p < 0.10$ , \*\* $p < 0.05$ , \*\*\* $p < 0.01$ .

**Table B.3:** Blinding Validation: Physician Identification of Assisted SOAP Note

| Panel A: Distribution of all responses |  |  |  |  |
| --- | --- | --- | --- | --- |
|  | Correct (%) | Incorrect (%) | Unsure (%) | N |
| All patients | 49.5 | 35.6 | 14.9 | 618 |
| Fully randomized sample | 50.4 | 36.2 | 13.3 | 450 |
| Panel B: Excluding unsure responses |  |  |  |  |
|  | Correct (%) | 95% CI | p-value | N |
| All patients | 58.2 | [53.9, 62.3] | 0.000 | 526 |
| Fully randomized sample | 58.2 | [53.3, 63.0] | 0.001 | 390 |

Physicians evaluated pairs of SOAP notes and were asked to identify which note was LLM-assisted. The answer options included "unsure" (to allow for abstention e.g. in the case of identical notes). The full sample includes all assessments with complete data. The fully randomized sample restricts to assessments after February 25, 2025. 95% confidence intervals use the Wilson score method. $p$ -values are from two-tailed binomial tests against 50% (chance performance).

**Table B.4:** Physician Assessment of Changes in Quality of Care, Controlling for Assisted Note Guess

|  | Qualitative error/harm |  |  |  | DALY loss |  |
| --- | --- | --- | --- | --- | --- | --- |
|  | Any error?* | Short-term harm | Long-term harm | Less harm | Severe loss* | Lower loss* |
| Effect of assisted note | -0.004<br>(0.016) | -0.010<br>(0.017) | -0.007<br>(0.005) | 0.006<br>(0.032) | -0.009*<br>(0.005) | 0.020<br>(0.026) |
| Physician guessed assisted | -0.038**<br>(0.018) | -0.054***<br>(0.020) | -0.015**<br>(0.006) | 0.158***<br>(0.037) | -0.012*<br>(0.006) | 0.109***<br>(0.030) |
| Order effect | 0.024<br>(0.016) | 0.014<br>(0.017) | -0.010*<br>(0.006) | -0.060*<br>(0.032) | -0.003<br>(0.006) | -0.011<br>(0.025) |
| Avg. unassisted note | 0.658***<br>(0.013) | 0.546***<br>(0.015) | 0.092***<br>(0.006) | 0.199***<br>(0.027) | 0.059***<br>(0.006) | 0.097***<br>(0.021) |
| R-squared | 0.02 | 0.02 | 0.03 | 0.06 | 0.02 | 0.04 |
| N (Patients) | 450 | 450 | 450 | 448 | 450 | 450 |

This table presents fixed effects regression estimates of the impact of AI-assisted clinical notes on various measures of medical assessment quality. The dependent variables include binary indicators for treatment errors and harm assessments (columns 1-4), as well as DALY (disability-adjusted life year) loss metrics (columns 5-6). The sample is restricted to assessments evaluated after February 25, when presentation order was randomized to control for order effect. Standard errors clustered at the patient level in parentheses. \* $p < 0.10$ , \*\* $p < 0.05$ , \*\*\* $p < 0.01$ .

**Table B.5:**
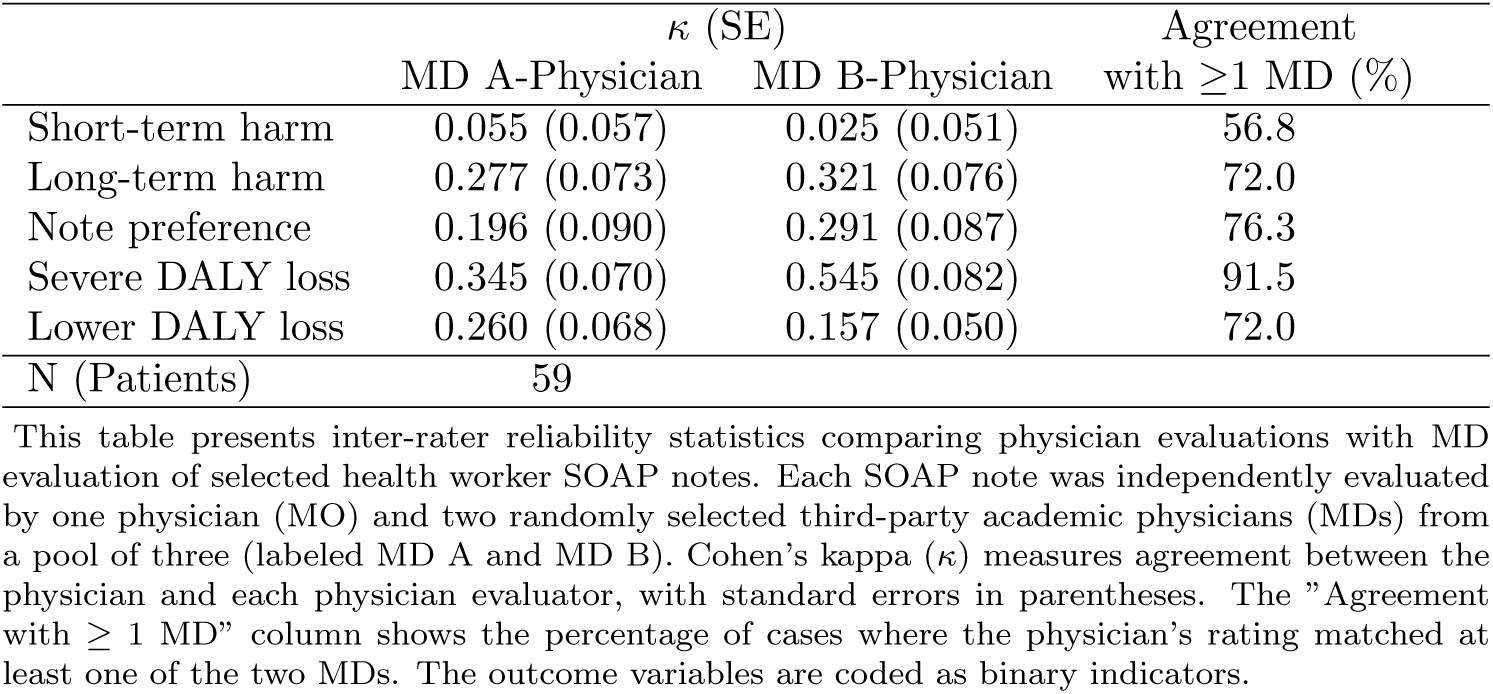
MD-Physician Rating Concordance

**Table B.6:**
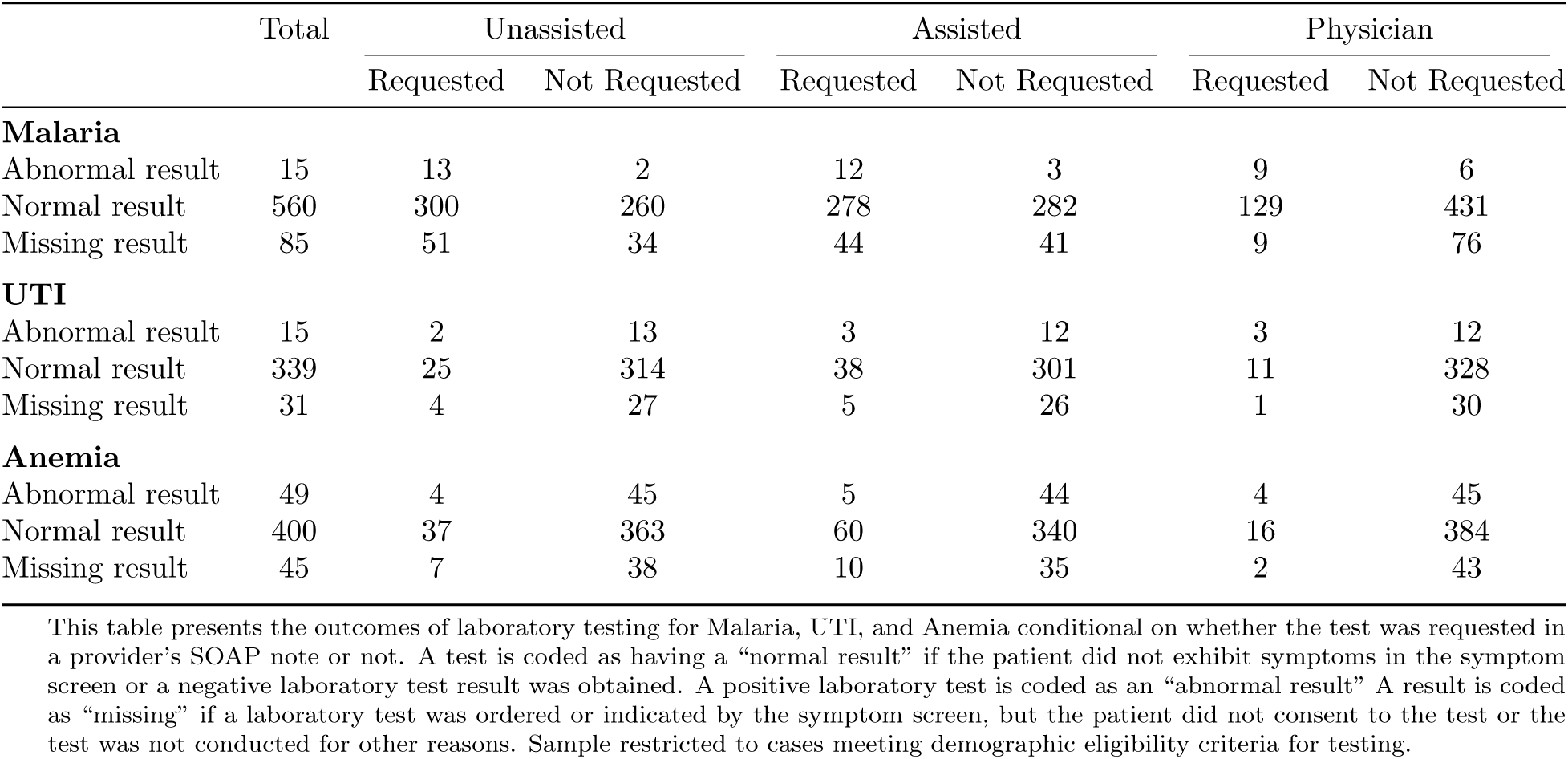
Laboratory Test Results by Provider Decision to Test in Unassisted, Assisted, and Physician SOAP Note

**Table B.7:** Tested Negatives and Missed Positives for Malaria, UTI, and Anemia (Tested Sample Only)

|  | Malaria |  | UTI |  | Anemia |  |
| --- | --- | --- | --- | --- | --- | --- |
|  | Tested<br>Negative<br>(1) | Missed<br>Positive<br>(2) | Tested<br>Negative<br>(3) | Missed<br>Positive<br>(4) | Tested<br>Negative<br>(5) | Missed<br>Positive<br>(6) |
| **Panel A** |  |  |  |  |  |  |
| Effect of assisted note | -0.034**<br>(0.016) | 0.003<br>(0.003) | 0.039*<br>(0.021) | -0.008<br>(0.008) | 0.052***<br>(0.014) | -0.003<br>(0.003) |
| Avg. unassigned note | 0.628***<br>(0.008) | 0.005***<br>(0.001) | 0.126***<br>(0.010) | 0.102***<br>(0.004) | 0.086***<br>(0.007) | 0.138***<br>(0.002) |
| R squared | 0.011 | 0.003 | 0.028 | 0.008 | 0.038 | 0.003 |
| N (Patients) | 382 | 382 | 127 | 127 | 327 | 327 |
| **Panel B** |  |  |  |  |  |  |
| Avg. assisted note | 0.594 | 0.008 | 0.165 | 0.094 | 0.138 | 0.135 |
| Avg. physician | 0.327 | 0.016 | 0.087 | 0.094 | 0.049 | 0.138 |
| Absolute testing rate (unassisted) | 0.662 |  | 0.142 |  | 0.098 |  |
| Absolute testing rate (assisted) | 0.626 |  | 0.189 |  | 0.153 |  |
| Absolute testing rate (physician) | 0.351 |  | 0.110 |  | 0.061 |  |
| Share abnormal test | 0.039 |  | 0.118 |  | 0.150 |  |

The outcome variables labeled “tested negative” are coded as 1 whenever the test was ordered by the health worker but the patient exhibited no related symptoms in the symptom screen or had a negative laboratory test. The variables for labeled “missed positive” are coded as 1 whenever the test was not ordered by the health worker but the patient had a positive test result either in the tests ordered as part of the study or in a test ordered by the physician.
Patient fixed effects included. Standard errors clustered at the patient level in parentheses. \* $p < 0.10$ , \*\* $p < 0.05$ , \*\*\* $p < 0.01$ .

**Table B.8:** Over-Ordering and Under-Ordering of Laboratory Tests for Malaria, UTI, and Anemia Relative to Physician Testing Decisions

|  | Malaria |  | UTI |  | Anemia |  |
| --- | --- | --- | --- | --- | --- | --- |
|  | Over<br>(1) | Under<br>(2) | Over<br>(3) | Under<br>(4) | Over<br>(5) | Under<br>(6) |
| Effect of assisted note | -0.044***<br>(0.011) | 0.002<br>(0.007) | 0.033***<br>(0.009) | -0.006<br>(0.004) | 0.058***<br>(0.011) | -0.008**<br>(0.003) |
| Avg. unassisted note | 0.358***<br>(0.005) | 0.029***<br>(0.003) | 0.062***<br>(0.005) | 0.027***<br>(0.002) | 0.095***<br>(0.005) | 0.036***<br>(0.002) |
| R squared | 0.025 | 0.000 | 0.020 | 0.004 | 0.042 | 0.008 |
| Patients | 660 | 660 | 660 | 660 | 660 | 660 |
| Share tested by MO | 0.223 | 0.223 | 0.045 | 0.045 | 0.045 | 0.045 |

Over-Ordering is defined as ordering a test the physician did not order. Under-Ordering is defined as not ordering a test the physician did order. Patient fixed effects included. Standard errors clustered at the patient level in parentheses. \* $p < 0.10$ , \*\* $p < 0.05$ , \*\*\* $p < 0.01$ .

**Table B.9:** Tested Negatives and Missed Positives for Malaria, UTI and Anemia (Fully Randomized Sample Only)

|  | Malaria |  | UTI |  | Anemia |  |
| --- | --- | --- | --- | --- | --- | --- |
|  | Tested<br>Negative<br>(1) | Missed<br>Positive<br>(2) | Tested<br>Negative<br>(3) | Missed<br>Positive<br>(4) | Tested<br>Negative<br>(5) | Missed<br>Positive<br>(6) |
| **Panel A** |  |  |  |  |  |  |
| Effect of assisted note | -0.014<br>(0.014) | 0.002<br>(0.002) | 0.030**<br>(0.013) | 0.000***<br>(0.000) | 0.059***<br>(0.014) | -0.003<br>(0.003) |
| Avg. unassisted note | 0.479***<br>(0.007) | 0.002**<br>(0.001) | 0.064***<br>(0.007) | 0.027***<br>(0.000) | 0.081***<br>(0.007) | 0.109***<br>(0.002) |
| R squared | 0.002 | 0.002 | 0.020 | . | 0.054 | 0.003 |
| N (Patients) | 424 | 424 | 264 | 264 | 321 | 321 |
| **Panel B** |  |  |  |  |  |  |
| Avg. assisted note | 0.465 | 0.005 | 0.095 | 0.027 | 0.140 | 0.106 |
| Avg. physician | 0.196 | 0.012 | 0.015 | 0.019 | 0.040 | 0.109 |
| Absolute testing rate (unassisted) | 0.505 |  | 0.064 |  | 0.090 |  |
| Absolute testing rate (assisted) | 0.488 |  | 0.095 |  | 0.153 |  |
| Absolute testing rate (physician) | 0.212 |  | 0.023 |  | 0.050 |  |
| Share abnormal test | 0.028 |  | 0.027 |  | 0.118 |  |

The outcome variables labeled “tested negative” are coded as 1 whenever the test was ordered by the health worker but the patient exhibited no related symptoms in the symptom screen or had a negative laboratory test. The variables for labeled “missed positive” are coded as 1 whenever the test was not ordered by the health worker but the patient had a positive test result either in the tests ordered as part of the study or in a test ordered by the physician.
Patient fixed effects included. Standard errors clustered at the patient level in parentheses. \* $p < 0.10$ , \*\* $p < 0.05$ , \*\*\* $p < 0.01$ .

**Table B.10:** Treatment Misallocation for Malaria, UTI and Anemia (Fully Randomized Sample Only)

|  | Overall* | Malaria | UTI | Anemia |
| --- | --- | --- | --- | --- |
| Effect of assisted note | -0.007<br>(0.004) | -0.007<br>(0.007) | -0.004<br>(0.004) | -0.009<br>(0.009) |
| Avg. unassisted note | 0.079***<br>(0.002) | 0.036***<br>(0.003) | 0.047***<br>(0.002) | 0.163***<br>(0.005) |
| R squared | 0.003 | 0.002 | 0.004 | 0.003 |
| N (Patients) | 1,040 | 447 | 267 | 326 |

Misallocation is coded as 1 when there is an intent to treat not conditioned on a positive test but the test is negative; or when the incorrect drug or dosage was prescribed or no treatment was prescribed and the test is positive. When the test result is missing, misallocation is coded as 1 when there is an unconditional incorrect drug or dosage, and 0 when a conditional plan specifies correct drug and dosage and otherwise as missing. Overall misallocation is prespecified in the pre-registration plan and at the patient-test level. Patient fixed effects included. Standard errors clustered at the patient level in parentheses. \* $p < 0.10$ , \*\* $p < 0.05$ , \*\*\* $p < 0.01$ .

**Table B.11:** Health Worker-Physician Objective Discordance (Count)

|  | Prescriptions |  | Diagnoses |  |
| --- | --- | --- | --- | --- |
|  | Over | Under | Over | Under |
| Effect of assisted note | 0.047**<br>(0.023) | -0.012<br>(0.014) | -0.002<br>(0.013) | 0.014<br>(0.010) |
| Avg. unassisted note | 1.683***<br>(0.012) | 1.058***<br>(0.007) | 0.802***<br>(0.007) | 0.974***<br>(0.005) |
| R-squared | 0.006 | 0.001 | 0.000 | 0.003 |
| N (Patients) | 660 | 660 | 660 | 660 |

This table presents fixed effects regressions examining the effect of LLM assistance on discordance between health workers and physicians. The dependent variables measure the number of mismatched items per patient assessment. "Over" indicates items documented by the health worker but not by the physician, while "Under" indicates items documented by the physician but not by the health worker. All models include assessment fixed effects and report robust standard errors. All notes in the study are included in the sample. Standard errors clustered at the patient level in parentheses. \* $p < 0.10$ , \*\* $p < 0.05$ , \*\*\* $p < 0.01$ .

**Table B.12:** Health Worker-Physician Objective Discordance (Fully Randomized Sample Only)

|  | Prescriptions |  | Diagnoses |  |
| --- | --- | --- | --- | --- |
|  | Any Over | Any Under | Any Over | Any Under |
| Effect of assisted note | -0.010<br>(0.011) | -0.010<br>(0.008) | 0.002<br>(0.011) | 0.000<br>(0.008) |
| Avg. unassisted note | 0.811***<br>(0.006) | 0.644***<br>(0.004) | 0.625***<br>(0.005) | 0.731***<br>(0.004) |
| R-squared | 0.002 | 0.003 | 0.000 | . |
| N (Patients) | 491 | 491 | 491 | 491 |

This table presents fixed-effects regressions examining the effect of LLM assistance on discordance between health workers and physicians "Any Over" is a dummy for whether there were any items documented by the health worker but not by the physician, "Any Under" indicates any items documented by the physician but not by the health worker. All notes in the study are included in the sample. Standard errors clustered at the patient level in parentheses. \* $p < 0.10$ , \*\* $p < 0.05$ , \*\*\* $p < 0.01$ .

## C Construction of Outcome Measures

### C.1 Subjective Physician Evaluation

Here we describe in detail how the various outcome variables are defined and constructed.

#### Any Treatment Error

During SOAP note evaluation, the physician indicates whether the note contains any error. The introductory text (here for SOAP Note A) is:

Treatment in SOAP Note A:

Please evaluate whether the treatment in SOAP Note A is appropriate for this patient’s condition. Please base this on *your own diagnosis*, not the health worker’s diagnosis in SOAP Note A. The treatment includes medications as well as other care instructions, such as information on home care or referrals.

This is followed by the question:

Is the treatment plan for the patient in SOAP Note A completely appropriate given your *own* diagnosis (accounting for conditional treatments based on medical tests)?

*Answer “No” if the patient should receive different medical care given your diagnosis. This can include both minor differences (for example, the patient should be advised to rest) and major errors (for example, the patient should receive a completely different set of medications)*.

(Answer options: yes/no/unsure)

We code this question as an indicator variable that equals 1 if the answer is “no”. If the answer is “unsure” the variable is treated as missing.

#### Deviations from the physician’s SOAP

The physician is also asked to assess for each SOAP note whether medical tests ordered were necessary or clinically useful vs. unlikely to be clinically useful. We code a variable that counts the number of tests that were unlikely to be clinically useful.

In addition, there are variables that ask whether there are missing or incorrect/unnecessary diagnoses, missing or incorrect/unnecessary medications, and missing or incorrect/ unnecessary care instructions. We code 6 indicator variables for any of these mistakes occurring in the SOAP note.

#### Short-term harm

To obtain a subjective assessment of patient harm, we ask the physician Now suppose the patient were to receive the treatment prescribed in SOAP Note (A) instead of your treatment. Please characterize what the consequences for the patient would be based on your *own* diagnosis.

What would be the short-term (temporary) consequences, such as additional symptoms or discomfort, that the patient would experience?

The answer options for this question are

- Severe: at least 7 additional days of being unable to carry out usual daily activities
- Moderately severe: 3-6 additional days of being unable to carry out usual daily activities
- Moderate: 1-2 additional days of being unable to carry out usual daily activities
- Mild: 2 or more additional days of discomfort, but no effect on usual daily activities
- None or minimal

#### Long-term harm

This question is followed by

Suppose the patient received the treatment in SOAP Note (A) instead of your treatment. Would this increase the risk of serious long-term (permanent) harm for the patient, including death?

with answer options yes, no, and unsure. We code an indicator variable that is 1 if the answer is yes and 0 otherwise. We also asked a follow-up question on the likelihood of such harm, but we do not show the results, because physicians gave identical likelihood ratings and in all other cases only one note received a likelihood rating, so this question does not convey additional information.

### C.2 Protocols for Malaria, UTI, and Anemia Testing

Within budget constraints, we aimed to test all relevant patients for malaria, UTI and anemia. We first specified a set of demographic inclusion criteria for participation in screening. All patients who fulfilled these criteria were considered part of the screening sample.

The testing itself consisted of a symptom screen followed by confirmation with a laboratory test. We coded the final result of the testing as either “normal” or “abnormal”. Patients with an abnormal test for the condition should generally receive treatment.

The symptom screen was designed to identify all patients who would plausibly be treated for the condition if they had a positive lab test. Patients who did not exhibit any relevant symptoms and did not have a test result from a physician-ordered test were coded as having a “normal” screening result. All patients who exhibited at least one relevant symptom and for whom the test was not already ordered by the physician were asked for their consent to conduct a laboratory test. For any patients for whom a test result was available (either test ordered by the physician, or passed the symptom screen and consented to testing), the screening was coded as “abnormal” with a positive test result and “normal with a negative test.

We applied the following demographic and symptom screening criteria:

#### Malaria RDT

All demographics are eligible. Malaria testing is offered if the patient reports a fever in the last 24 hours.

#### Anemia/PCV test

All patients 18 years and older. Anemia testing was limited to adults due to the need to collect a venous blood sample, which is more invasive and technically difficult in children. The test is offered for patients reporting feeling tired or with low energy for 4 weeks or more, feeling like their heart is racing for 4 weeks or more, signs of bleeding for 4 weeks or more (vaginal bleeding, blood in stool or in urine), feeling dizzy, difficulty breathing, currently pregnant, and delivered a baby within the past six weeks.

#### Urine dipstick analysis

Only female patients 7 years or older. This test was restricted to females given the substantially lower prevalence of urinary tract infections in males and finite study resources. The lower age limit of 7 years reflects the practical requirement for patients to provide a urine sample on request. Urine dipstick analysis is offered for patients reporting pain while urinating, blood in urine, frequent urge to urinate, abnormal discharge from the genital area, and burning feeling while urinating.

### C.3 Misallocation of Treatment for Malaria, Anemia and UTI

In order to determine whether a patient received the correct treatment for the three screening conditions, we proceeded in multiple steps.

First, we determined using the clinical indication, test condition attached to a prescription, or drug itself whether the SOAP note indicated an intent to treat for the condition.

Second, we used the treatment guidelines below to determine whether the provider specified the correct treatment for the condition they intended to treat.

Finally, we used information in the conditioning field for each prescription to determine whether they intended to treat the patient only with a positive laboratory test result or unconditionally.

We then coded as “overtreatment” any case in which the provider intended to treat for a condition, did not specify that the treatment was conditional on a test result, and the screening test or a physician-test was normal/negative. This approach accounts for giving the patient a drug intended for the condition in question but choosing a incorrect dosage or treatment option. For example, some health workers prescribe vitamins for anemia but choose a product that does not contain iron; if the patient’s PCV test is normal, this is overtreatment because it involves an unnecessary medication.

We code as “undertreatment” any case in which the provider did not intend to treat for the condition, or intended to treat but prescribed an incorrect drug, and the screening test or a physician-test was abnormal/positive. This accounts for cases where the provider meant to treat but gave something incorrect. For example, a vitamin prescription that does not contain iron is undertreatment for anemia because it is missing the relevant ingredient to address the condition.

Note that we define misallocation at the level of the condition, not the level of the patient. A patient who receives the wrong treatment because they are misdiagnosed is overtreated for one condition and undertreated for another. We only measure misallocation for three conditions and so we cannot fully assess misallocation at the patient level (i.e., “was this patient treated for the condition they actually have?”).

The treatment guidelines we applied are:

- For malaria:

**Adults:** Artemether-Lumefantrine 80 mg/480 mg by mouth twice daily for 3 days

**Children 5-14 Kg:** Artemether-Lumefantrine 20 mg/120 mg by mouth twice daily for 3 days

**Children 15-24 Kg:** Artemether-Lumefantrine 40 mg/240 mg by mouth twice daily for 3 days

**Children 25-34 Kg:** Artemether-Lumefantrine 60 mg/360 mg by mouth twice daily for 3 days

**Severe malaria:** Artemether 3.2 mg/Kg IM loading dose once followed by 1.6 mg/kg IM maintenance dose once daily for 3 days

- For anemia:

**Adults:** Ferrous Sulfate 200 mg by mouth once daily for one month

**Children:** Ferrous Sulfate 125mg/ml syrup 3-6mg/kg/day for one month

**Pregnant women:** Ferrous Sulfate 200 mg by mouth twice or thrice daily for one month

**Severe anemia:** Refer for transfusion

- For UTI (uncomplicated):

**Adults:** first-line treatment Nitrofurantoin 100 mg by mouth twice daily for 7 days OR second-line treatment Cotrimoxazole 960 mg by mouth twice daily for 5 days

**Children:** Cotrimoxazole 8-10 mg/Kg/day by mouth divided twice daily for 5 days

**Pregnant women in third trimester only:**

Amoxicillin 500 mg by mouth every 8 hours for 7 days

OR

Amoxicillin/Clavulanate 500/125 mg by mouth twice daily for 3-7 days

OR

Cephalexin 500 mg by mouth twice daily for 7 days

### C.4 DALY Harm Scales

To measure the expected healthy time lost translated to DALY, we used a novel multi-step survey procedure combined with extensive training about how to evaluate harms. Iatrogenic health harms from care plan errors can span many orders of magnitude, from a temporary, minor symptom to permanent disability or death.

Therefore, our approach entailed an anchored scale that provided ordered benchmark errors relevant to local clinical practice. Treating physicians first made an ordinal determination regarding the harm magnitude of the care plan they were grading, relative to the benchmark errors. Then, the treating physicians selected a harm magnitude along a visual analog scale linking these reference errors. Because we calculated DALY estimates for health losses due to each reference error, this approach allowed us to numerically capture physicians’ judgment of health plan harms without any explicit calculations involving the care plan’s effects on the likelihood of experiencing various health states, the expected duration of those health states, or the reduction in health experienced by patients in each health state.

After being guided through a process of detailing qualitative differences between the health worker’s note and their own, the treating physicians were asked to specify the temporary and permanent harms from implementing the health worker’s care plan rather than their own. The treating physicians were presented with the following survey prompt:

“Consider the overall health harms from the treatment errors in SOAP Note [A/B] that you just described (temporary symptoms and risk of longterm or permanent harm).

The scale below shows levels of harm that average [adults/children] experience due to different medical errors, ordered from least to most severe.

Harm is expressed in terms of ‘healthy time lost’.

*For example, unpleasant symptoms for 6 days reduce quality of life by 1/3 for those days, resulting in the equivalent of 2 days of life lost*.

Please select the appropriate interval of the harm scale for SOAP Note [A/B].”

Below this prompt, the scale of different levels of harm is shown in table form, as in figure B.1.

The physician uses the multiple-choice response option in the bottom to select an interval on the scale.

On the next screen, they see a detailed description of the two scale end-points and use a slider to choose the estimated level of harm between those two points, see Figure B.2 for an example.

The DALY scale has 9 benchmarks from 0 minutes to 23 years (28 years for children), generating 8 “bins” each. In detail, the benchmarks are shown in Table B.13.

#### Estimating DALY harms for benchmark errors

DALY estimates for benchmark errors were quantified by physician members of our research team. Research within the field of health decision science tends to employ calibrated simulation models to quantify the DALY or QALY health effects of alternative health interventions (Hunink et al., 2014). Such simulations require inputs such as utility weights for relevant health states, the expected duration of each health state for patients, and transition probabilities between health states, which are affected by the health care interventions being studied. Although these studies tend to compare alternative treatment protocols or public health interventions, rather than examining specific clinical errors, such research and related clinical effectiveness research contain estimates that can be used to approximate the DALY effects of health care errors.

**Table B.13:**
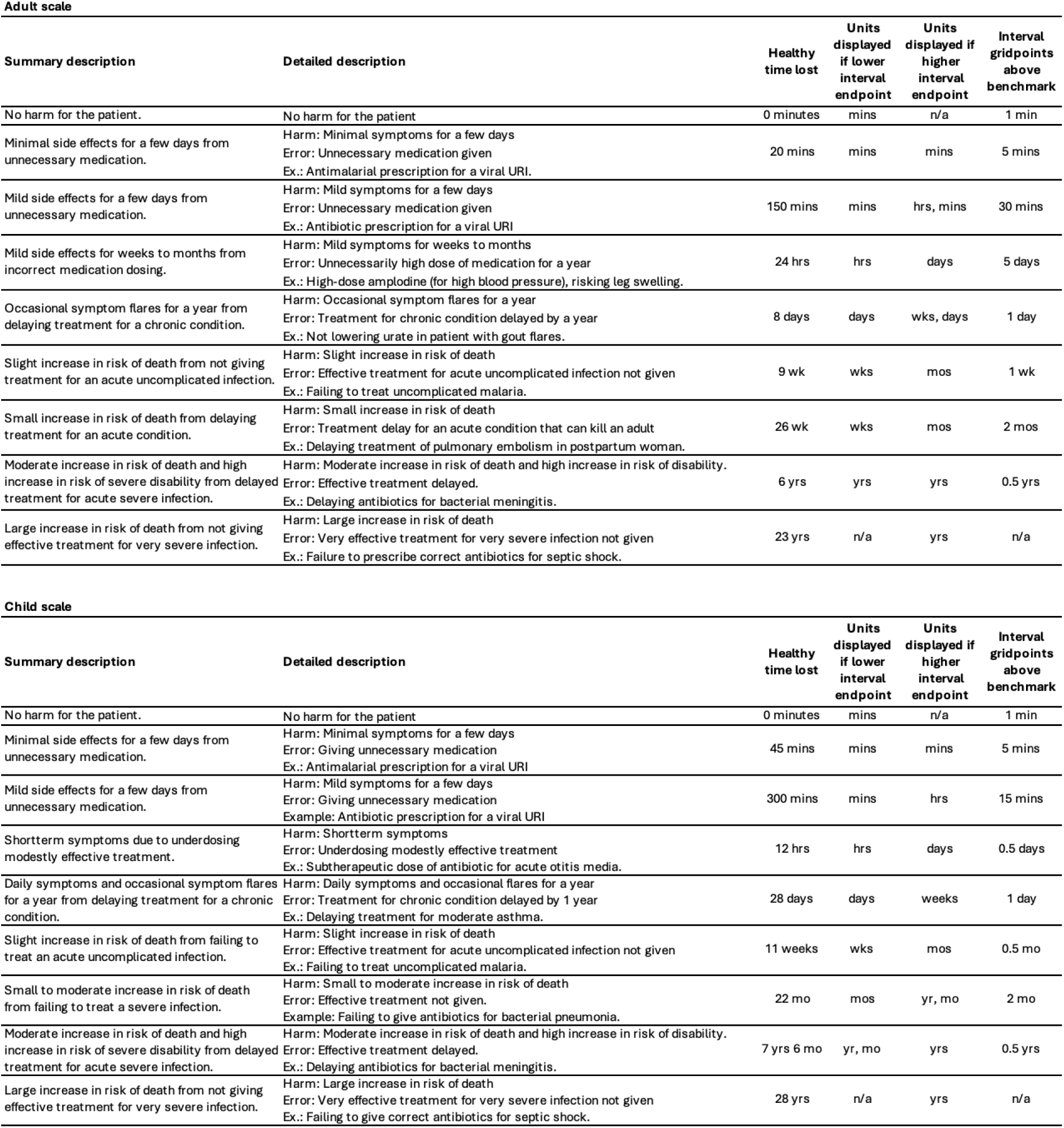
Adult and child scale harm benchmarks.

**Figure B.1:**
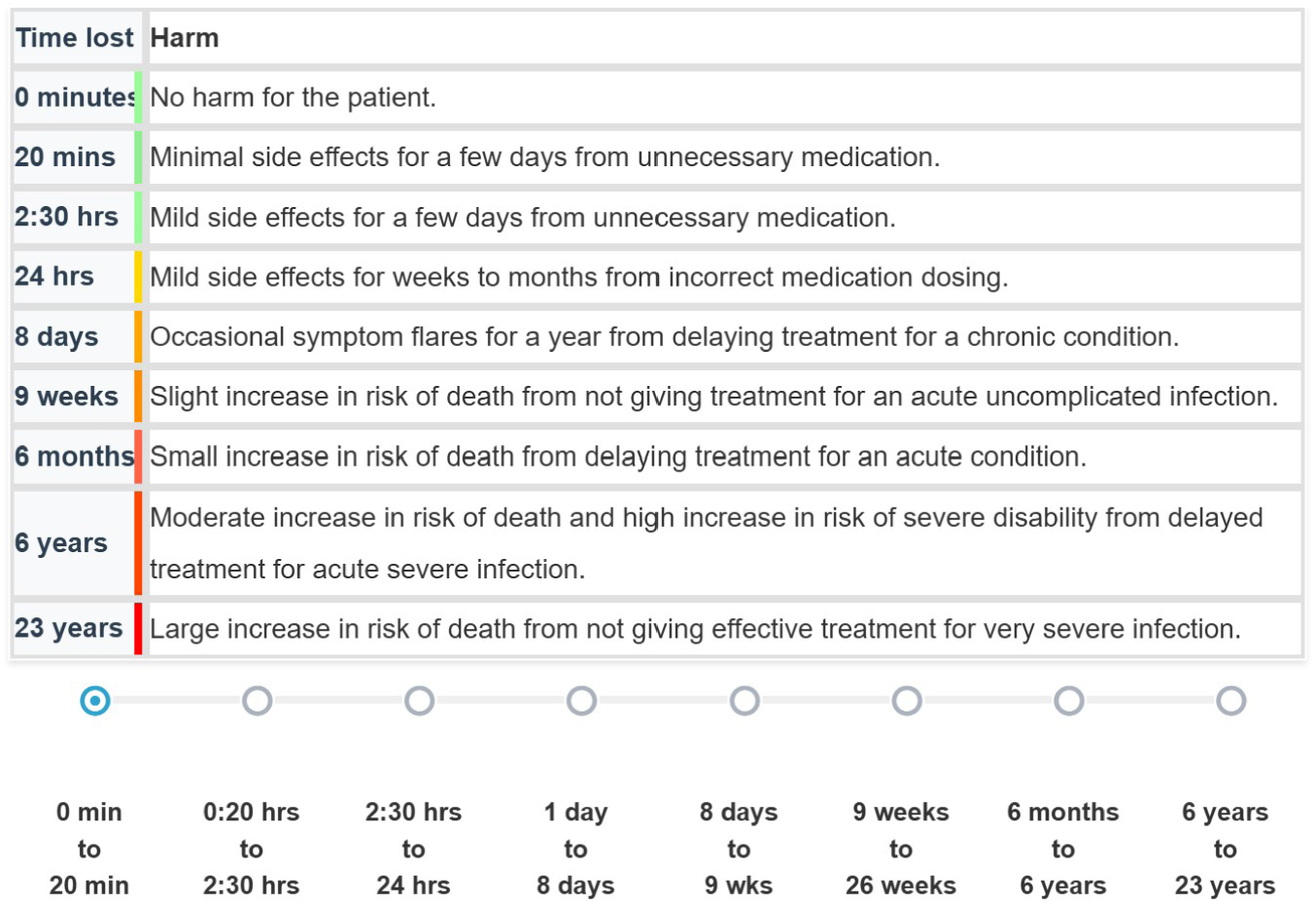
Initial Rank Ordering (Adult Scale)

**Figure B.2:**
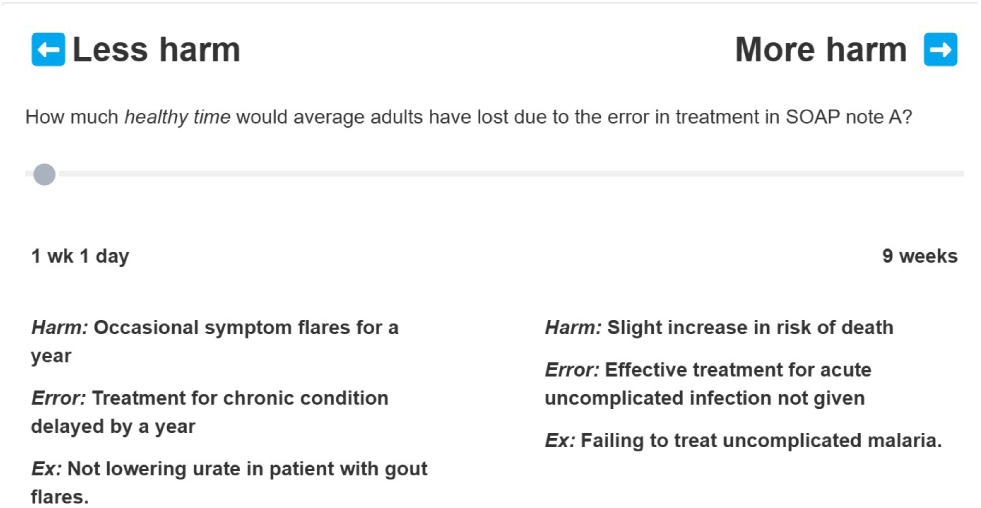
Example of Healthy Time Loss Quantification

DALY harms of some benchmark errors are drawn directly from prior studies. For example, our estimate of the DALY harm of failing to treat a child for uncomplicated malaria, 11 weeks of healthy time lost, is the difference between the published DALY estimates of treated versus untreated uncomplicated malaria for a 5 year old in sub-Saharan Africa (Basu et al., 2014). Similarly, our estimate of the DALY harm of failing to treat gout with urate-lowering medications, 8 days of healthy time lost, was drawn directly from a prior simulation study (van de Laar et al., 2022).

For other benchmark errors, DALY estimates are derived from simple calculations drawing from multiple prior studies. For example, our estimate of healthy days lost from an unnecessary pediatric antibiotic prescription, 5 hours, reflects the estimated incidence of antibiotic-associated diarrhea (23%) and its mean duration (4 days), both from Turck et al. (2003), as well as estimates for the utility losses for moderate diarrhea (18.8%) and daily medication use/worry (4.9%), both from Salomon et al. (2015). For the most severe benchmark errors, DALY estimates reflect expected healthy years of life lost from the increased likelihood of mortality. For example, for the benchmark error of failing to give correct antibiotics for a child with septic shock, the calculated DALY estimate of healthy life lost is 28 years. This estimate reflects the 5.05-fold lower mortality observed when patients with septic shock receive appropriate antibiotics (Kumar et al., 2009), a 12.2% baseline 2-day mortality rate for children with malaria-negative sepsis in sub-Saharan Africa (Maitland et al., 2011), and the remaining life expectancy for 5 year-old children in Nigeria (65.67 years), which we convert to expected healthy life years remaining (57.05) based on the ratio of Nigeria’s life expectancy at birth to its healthy life expectancy at birth (World Health Organization, 2025).

#### Severe error

Because the distribution of DALY harms was expected to be heavily skewed, with a large proportion of zeros, we also create binary indicators for the presence of a severe error. We define severe errors as any errors at or above the 95th percentile of DALY harms for unassisted care plans (pooling the harm ratings for children and adult).

#### An indicator for the better care plan

Based on the DALY rating of SOAP Note A vs. B (counting instances with no errors as 0 DALY loss) we will define an indicator that is 1 if the SOAP Note has strictly lower DALY loss. This indicator is 0 for both notes if the notes are judged to be the same according to this question, asked after the physician rated Note A:

Are there any *meaningful* differences in the treatment plans of SOAP Note A and B?

Only if the answer is “yes” or “unsure”, the review of SOAP Note B proceeds.

## Footnotes

1 CHEWs at EHA Clinics are able to order and interpret results of some basic laboratory tests.

2 Due to the difference in serious illness rates between our two samples, we replicate results from the larger sample shown below in the smaller sample and confirm that they are qualitatively unchanged (Appendix Tables B.9 B.10, and B.12, see also sections 3.3 and 3.4).

3 We did not record whether the physician thought any medical tests were missing. CHEWs are authorized to request only a small number of tests that can be 1:1 compared with the MO, and we do so in section 3.4. The question on medical tests asked instead whether a given test was “necessary”, “not necessary but clinically useful” or “unlikely to be clinically useful”. This was based on findings from prior pilots that CHEWs tend to use laboratory testing more than MOs, and allows for the possibility that CHEWs need to use medical tests for diagnosis where MOs use their greater diagnostic skills.

4 While false negatives are rare, laboratory tests can produce false positives. We here assume that the reported symptoms which qualified patients for testing, combined with the test result, provide sufficient likelihood of the condition to warrant treatment.

5 Exact counts of false negatives, false positives, true negatives and true positives for the assisted and unassisted notes for each condition are shown in Table B.6.

6 The study took place in the dry season when malaria rates are low.

7 Some researchers have argued for more universal testing on the basis of low testing costs, although this departs from the usual standard of care in our setting (Shillcutt et al., 2008).

8 We define misallocation at the level of the condition. At the level of the patient, misdiagnosis means that overtreatment for one condition often coincides with undertreatment for another. Note also that empiric therapy can be the correct approach when diagnostic tools are unavailable or there is a life-threatening emergency. This would not be the case for patients in this study.

9 Two PIs reviewed all diagnoses in the study and all treatments for these diagnoses blinded to their origin SOAP note and created the groupings. This approach was intended to account for different forms of giving the same medication (e.g. as a syrup or a pill) and different brand names, along with idiosyncratic differences in the use of the ICD-10 based diagnosis options, which were unlikely to be clinically important. We use the conditional physician SOAP notes for comparability. For the diagnoses, we use either the “clinical indication” fields or the main “diagnosis” field in the EMR if there are no prescription drugs in the SOAP note.

10 The first steps above were done within a single prompt that evaluated the health worker’s unassisted note, the LLM feedback, and the physician’s note. This last step was completed with a prompt that evaluated the structured list of annotated recommendations and the health worker’s assisted and unassisted notes.

